# Estimating the Efficacy of Common Treatments in Children and Young Adults Diagnosed with Cerebral Palsy Using Three Machine Learning Algorithms

**DOI:** 10.1101/2021.10.06.21264624

**Authors:** Michael H. Schwartz, Andrew J. Ries, Andrew G. Georgiadis

**Affiliations:** Gillette Children’s Specialty Healthcare; University of Minnesota Department of Orthopedic Surgery

## Abstract

**Background:** Orthopedic and neurological deformity are often treated in children and young adults with cerebral palsy (CP). Due to challenges arising from combinatorics, research funding priorities, and medical practicalities, the efficacy of these treatments is not well studied.

**Objectives:** Our goal was to estimate the efficacy of 13 common orthopedic and neurological treatments at four different levels of outcome in children and young adults diagnosed with CP. The outcome levels considered were anatomy and physiology, gait parameter, overall gait pattern, and function.

**Methods:** We used three well-establish causal inference approaches (direct matching, virtual twins, and Bayesian causal forests) and a large clinical gait analysis database to estimate the average treatment effect on the treated (ATT). We then examined the efficacy across treatments, methods, and outcome levels.

**Results:** The median ATT of 13 common treatments in children and young adults with CP, measured as Cohen’s D, bordered on medium at the anatomy and physiology level (median [IQR] = 0.42 [.05, .60]) and became smaller as we moved along the causal chain through gait parameter (0.21 [.01, .33]), overall gait pattern (0.09 [.03, .19]), and function (−0.01 [-.06, .13]).

**Conclusions:** Current treatments have medium effects on anatomy and physiology, but modest to minimal efficacy on gait and function. Further work is needed to understand the source of heterogeneous treatment effects, which are large in this patient population. Replication or refutation of these findings by other centers will be valuable to establish the generalizability of these results and for benchmarking of best practices.

## 1 Background

### 1.1 Cerebral palsy and Orthopedic Deformity

There are approximately 750,000 people in the United States currently diagnosed with cerebral palsy (CP), and 10,000 new cases each year (1–3). Medical costs for children diagnosed with CP are 10 to 26 times higher than for nondisabled children (4). Around 70% of individuals diagnosed with CP walk independently or with walking aids (59% and 11%, respectively) (5). The primary neurological impairments commonly found in individuals with CP include spasticity, reduced motor control, and weakness. Over time, these neurological impairments often lead to orthopedic deformity.

Neurological and orthopedic problems are frequently treated by surgery or neurotoxin injections. The rationale for treatment is that deformity impairs gait and mobility, which impairs function and reduces quality of life. Thus, by intervening at the deformity level, it is hoped that changes will propagate through the causal chain, ultimately leading to improvements in function and quality of life. Accordingly, treatment goals are usually multi-level, including (1) anatomical and physiological goals, such as reducing excessive femoral anteversion or reducing spasticity, (2) specific gait goals, such as correcting intoeing, (3) overall gait goals, such as improving the walking pattern, and (4) functional goals, such as improving performance on mobility-related tasks like stair climbing.

Because problems don’t occur in isolation, surgery in children and young adults with CP is often executed at multiple levels during a single operation (single-event multi-level surgery – SEMLS). This makes it hard to estimate the isolated impact of an individual surgery. For one thing, the 13 relatively common surgeries considered in this study can be combined in 8191 unique ways. Furthermore, there is limited funding available to study the effectiveness of established treatments in CP (6). Finally, given how well-established most of the treatments in CP are, it would be difficult to find patients and surgeons willing to participate in randomized controlled trials (RCTs). The absence of strong evidence means that most of what we know about treatment outcome in CP is based on observational studies. The design of these studies is often insufficient to establish strong evidence. Examples include not comparing to a control group, relying on case studies, and deferring to expert opinion (7).

### 1.2 Causal Inference

Observational studies are susceptible to selection bias. Patients receiving different treatments are not randomized, and thus differ in their characteristics. In addition, patients are generally chosen for treatments based on a doctor’s reasonable belief that the patient will either benefit from the treatment, fare poorly without a treatment, or both. This last element, known as targeted selection, causes important but often unrecognized problems when estimating treatment outcomes in observational studies (8,9). Despite these challenges, it is critical that we understand the effectiveness treatments for individuals with CP.

An RCT is the gold standard for establishing causal inference, but it is not the only option (10). There are many statistical and machine learning methods that can be used to estimate treatment effects (11). These methods rely on adjusting for and regressing on important covariates that determine both treatment assignment and treatment outcome. Causal inference has gained popularity over the years and has been validated by reproducing the results of RCTs and deriving accurate effects from synthetic data. For example, in the context of CP treatment, we have recently shown that a standard causal inference technique can accurately and precisely estimate the effects of rectus femoris transfer compared to an RCT (12). We have used similar methods to estimate the effect of SEMLS (13).

In the present study we will use three modern causal inference methods: direct matching (DM), virtual twins (VT), and Bayesian Causal Forests (BCF) to estimate average treatment effects on the treated (ATTs) for 13 common treatments in children and young adults with CP. We briefly describe these methods below. We will estimate outcomes at four levels: anatomy and physiology, gait parameter (i.e., specific deviations in gait kinematics), overall gait pattern, and function. The DM model will generate a matched subset of treated observations. We will then use this matched subset to estimate an ATT with both the VT and BCF models. We will also estimate an ATT from the VT and BCF models using the entire set of treated observations. By comparing the ATT estimates obtained from the matched subset to those obtained using all treated observations, we will identify possible bias due to omitted observations. We will consider the three models’ estimates together when interpreting the results. In doing so we will obtain a robust picture of the overall effectiveness of common treatments used for correcting deformity in children and young adults with CP.

## 2 Methods

This study was reviewed and authorized by the University of Minnesota institutional review board review (STUDY00012420).

All results from causal inference methods are conditional on modeling assumptions. In this study, we follow the principles of the Rubin causal inference framework (14,15). We assume that by controlling for the appropriate set of causal pre-treatment covariates, either through matching (DM) or modeling (VT, BCF), observational data can be used to estimate the causal average treatment effect. Choosing the proper set of covariates is, of course, the crucial decision in this approach, and will be described below.

### 2.1 Participants and Covariates

#### 2.1.1 Participants

We queried our database for individuals diagnosed with CP, less than 25 years old, who had two standard clinical gait assessments at least nine months and no more than 30 months apart. We considered limbs, rather than individuals, as observations. This was motivated by the asymmetry commonly observed in this patient population and the standard clinical process that generates treatment decisions based primarily on limb-level data. We used bootstrap confidence interval estimates to avoid making any assumptions about limb independence.

#### 2.1.2 Covariates

A uniform set of covariates was chosen for all predictive models. We used the same covariates to derive propensity score models for each treatment. The propensity scores were used as inputs to the predictive models, and are particularly important for the BCF approach. The covariates used in the models describe diagnosis, anthropometry, time and distance parameters, neurological impairments, contracture, bony alignment, kinematic gait deviations, and both prior and interval treatment [Table 1]. The covariates were chosen pragmatically to span the patient factors that are measured, analyzed, and discussed when devising a treatment plan. We also chose measures that are likely to be obtained at most clinical gait centers in order to promote future efforts to replicate or refute the findings presented here. The variable names are largely self-explanatory, but a complete glossary is provided (Appendix 1).

**Table 1.** Covariates in causal models.

| Category | Variables |
| --- | --- |
| Diagnosis | topographic sub-type, diagnosis side,<br>whether limb is affected or unaffected (unilateral sub-types) |
| Anthropometry | Age, sex |
| Time and Distance<br>Parameters | timing of foot-off, opposite foot-off, and opposite foot contact,<br>dimensionless speed and step length (17) |
| Neurological<br>Impairments | <u>spasticity</u>: Modified Ashworth Scores for hip adductors, hip flexors,<br>hamstrings, plantarflexors, and rectus femoris<br><u>strength</u>: Manual Muscle Test Grades for hip abductors, hip flexors and<br>extensors, knee flexors and extensors, and plantarflexors<br><u>static selective motor control</u>: Clinical grade of absent, diminished, or<br>typical control for hip abductors, hip flexors, hip extensors, knee<br>flexors, knee extensors, and ankle plantarflexors |
| Contracture | <u>ankle</u>: maximum passive ankle dorsiflexion (knee at 0° and 90° of<br>flexion)<br><u>knee</u>: maximum passive knee flexion and extension<br><u>hip</u>: maximum passive hip flexion and extension |
| Bony Alignment | <u>tibial torsion</u>: bimalleolar axis angle<br><u>femoral anteversion</u>: anteversion estimated by trochanteric prominence<br>test, maximum hip internal and external rotation |
| Kinematic Gait<br>Deviations | Gait Deviation Index<br>Discretized kinematic data for Levels × Planes × Measures, where,<br><u>levels</u> = pelvis, hip, knee, ankle, and foot,<br><u>planes</u> = sagittal, coronal, and transverse plane,<br><u>measures</u> = angle value at initial contact and foot-off, angle value<br>and timing of maximum and minimum during stance, swing, and<br>overall, mean angle, angle range of motion during stance, swing, and<br>gait cycle |
| Mobility-Related<br>Function | Gross Motor Function Classification System (GMFCS) level |
| Prior and Interval<br>Treatment | <u>neurological</u>: selective dorsal rhizotomy, botulinum toxin type A or<br>phenol injection, intrathecal baclofen pump implantation, other<br>neurosurgery (e.g., shunt placement, neurectomy)<br><u>bony</u>: femoral derotation osteotomy, tibial derotation osteotomy, foot<br>and ankle bony surgery, distal femoral extension osteotomy<br><u>soft tissue</u>: adductor release, foot and ankle soft tissue surgery, calf<br>muscle lengthening, psoas release, hamstrings lengthening, patellar<br>advance, rectus femoris transfer<br><u>casting</u>: lower leg cast (short or long)<br><u>propensity</u>: probability of receiving one of the 13 target treatments |

##### Gait and Clinical Examination Measures

Three-dimensional gait kinematics were measured at baseline and follow-up. Gait parameters were computed as the mean of three to five barefoot over-ground walking trials collected at a self-selected speed. Our motion analysis laboratory used modern, three-dimensional gait analysis equipment and methodology, and employed highly experienced staff. Kinematics were computed using a modification of the Vicon Plug-in-Gait model (Vicon Motion Systems Ltd, UK) with hip centers and knee axes identified using functional methods, and malleoli identified using virtual markers (16,17). Observations with knee varus-valgus range-of-motion > 15°were removed to enhance the quality of the transverse plane kinematic profile (18).

Physical examinations were performed by licensed physical therapists. Spasticity was scored using the modified Ashworth scale (19). Strength was estimated from a manual muscle test (20). Static selective motor control at various levels was graded as absent, diminished, or typical. Range-of-motion was assessed passively using a hand-held goniometer.

##### Function

Mobility-related function was measured using the Functional Assessment Questionnaire Transform (FAQt) (21). The FAQt is a difficulty-weighted average of the 23 mobility skills queried by the Functional Assessment Questionnaire (22).

##### Missing data

There are valid reasons to believe that some missing data occur in meaningful clinical patterns. For example, it is common to find missing data among neurological covariates (strength, spasticity, selective motor control) in individuals with significant cognitive impairments, due to the patient’s inability to understand and follow directions. These same impairments are correlated with overall severity and may also impact treatment outcome due the child’s ability to participate fully in rehabilitation after surgery. Missing values for the FAQt were imputed if ≥18/23 questions upon which the FAQt depends were present. The `mice` package in R was used for imputation of FAQt based on available FAQ skill values (23). Missing values in categorical data were assigned a value (“Miss”). This protects against data that are not missing (completely) at random.

##### Propensity scores

Propensity scores used in the DM, VT, and BCF models were computed from a separate BART model using the `bartMachine` package in R (24). Propensity score modeling is not the focus of this paper, and many good methods exist for estimating propensity scores (25). The propensity model performance on independent test set data is included for reference [Table 2].

**Table 2.** Performance of propensity score models.

| **Treatment** | **Acc.** | **Sens.** | **Spec.** | **AUC** |
| --- | --- | --- | --- | --- |
| Selective Dorsal Rhizotomy | 0.85 | 0.90 | 0.84 | 0.93 |
| Chemodenervation Injection | 0.62 | 0.54 | 0.65 | 0.64 |
| Rectus Transfer | 0.79 | 0.90 | 0.78 | 0.91 |
| Femoral Derotation Osteotomy | 0.77 | 0.76 | 0.78 | 0.85 |
| Tibial Derotation Osteotomy | 0.79 | 0.76 | 0.79 | 0.84 |
| Psoas Release | 0.76 | 0.84 | 0.75 | 0.87 |
| Adductor Release | 0.71 | 0.82 | 0.70 | 0.85 |
| Hams Lengthening | 0.76 | 0.80 | 0.76 | 0.84 |
| Calf Muscle Lengthening | 0.77 | 0.70 | 0.78 | 0.82 |
| Distal Femoral Extension Osteotomy | 0.88 | 0.96 | 0.88 | 0.98 |
| Patellar Advance | 0.85 | 0.91 | 0.85 | 0.94 |
| Foot and Ankle Bone | 0.68 | 0.81 | 0.65 | 0.80 |
| Foot and Ankle Soft Tissue | 0.68 | 0.68 | 0.68 | 0.73 |

Acc – accuracy, Sens – sensitivity, Spec – specificity, AUC – area under the receiver operation characteristic curve
Note: All results are for out-of-sample (independent) test data

### 2.2 Treatments and Outcomes

At our center, the 13 treatments we will focus on in this study account for over 93% of the treatments performed on children and young adults seen for pre- and postoperative three-dimensional gait assessment [Table 3]. These are consistent with the most common treatments performed in this population (26). We have defined relatively broad treatment categories. For example, the treatment category “*calf muscle lengthening*” groups together a variety of different surgical techniques, such as Baker and Strayer. Our coarse-grained approach is intended to emphasize the “*big picture*” nature of this study. Differences in outcomes between sub-categories within a given treatment category (e.g., Baker vs. Strayer) are not considered here. Note that interval treatment includes all treatment between baseline and follow-up gait analysis. Interval treatment usually, but not always, occurs at a single event. Treatments are recorded in our database based on the patient’s medical record. Most, but not all, treatments occurred at our center.

**Table 3.** Treatments and Outcome measures.

| **Surgery** | **Anatomy & Physiology** | **Gait Parameter** |
| --- | --- | --- |
| Selective Dorsal Rhizotomy | Mean Spasticity § (meanspas) | Mean stance ankle dorsiflexion |
| Neurotoxin Injection | Mean Spasticity | Mean stance ankle dorsiflexion |
| Femoral Derotational Osteotomy | Femoral Anteversion | foot progression deviation from typical (mean over stance) |
| Tibial Derotational Osteotomy | Bimalleolar axis angle deviation | foot progression deviation from typical (mean over stance) |
| Foot and Ankle Bone Surgery | Weight-Bearing Foot deformity severity † (footsev) | foot progression deviation from typical (mean over stance) |
| Distal Femoral Extension Osteotomy | Knee extension | Mean stance knee flexion |
| Psoas Release | Maximum Hip Extension | Minimum stance Hip Flexion |
| Hamstrings Lengthening | Popliteal angle | Minimum swing knee flexion |
| Adductor Lengthening | Hip abduction with knee extended | Mean stance hip abduction |
| Calf Muscle Lengthening | Ankle dorsiflexion with knee extended | Mean stance ankle dorsiflexion |
| Rectus Femoris Transfer | Rectus femoris spasticity ‡ | Maximum swing knee flexion |
| Patellar Advancement | Knee extensor lag | Mean stance knee flexion |
| Foot and Ankle Soft Tissue Surgery | Weight-Bearing Foot deformity severity | foot progression deviation from typical (mean over swing) |

§ Mean Spasticity (meanspas) = Ashworth score averaged over adductors, hamstrings, rectus femoris, plantarflexors
† Foot deformity severity (footsev) = numerical severity score (0-typical – 3-severe) averaged over weight-bearing hindfoot and forefoot severity assessment
‡ As measured by physical examination

Outcomes were assessed at four levels for each treatment: anatomy and physiology, specific gait parameter, overall gait pattern, and function. For all treatments, overall gait pattern was measured by the gait deviation index (GDI) and function was measured by the FAQt (21). Outcomes at the level of anatomy and physiology and specific gait parameter were chosen for each treatment using clinical experience [Table 3].

### 2.3 Models

All computations were performed in R (27). We used the `designmatch` package for the DM estimate, the `bartmachine` package for the VT estimate, and the `bcf` package for the BCF estimate (9,24,28).

#### Direct Matching (DM)

For the DM approach, treatment effects are estimated from the difference in outcome between one-to-one matched treated and control observations. Matched controls are obtained by imposing the following constraints:

- **Distance**. Minimize the multivariate distance (Mahalanobis rank distance) between treated and control observation based on a set of relevant, treatment-specific physical examination and gait kinematic parameters. Penalize mismatches of propensity score (the probability of an observation undergoing a treatment, given a set of covariates) when they exceed a standardized mean difference of 0.2. This “caliper” on propensity score ensures that we match both the covariates and the propensity score.
- **Near-Fine Balance**. Match the groupwise distributions of treatments and, for certain treatments, key categorical physical examination measures that are not well balanced with distance matching alone.
- **Moment Balance**. Match the means of relevant physical examination measures and gait kinematic parameters on a groupwise basis (treated vs. control).

For the DM estimate we used the `bmatch` function and the optimal subset approach, with the subset weight set to the median of the distance matrix and the `glpk` solver to find an approximate solution.

#### Virtual Twins (VT)

For the VT approach we first build a predictive model of the outcome. Next, we generate a fabricated counterfactual version (virtual twin) of each observation. For example, if the observation was treated, the virtual twin is created by setting the treatment status to untreated while leaving all other covariates unchanged. An outcome prediction is then made on the virtual twin, and the treatment effect is computed as the difference between the actual and virtual twin outcomes. In our implementation of the VT estimate, we use Bayesian Additive Regression Trees (BART) as the predictive model using the `bartMachine` function with all default settings.

#### Bayesian Causal Forests (BCF)

In the BCF approach we use an underlying BART model, but in a manner substantially different from the VT approach. A BCF is a modification of the traditional BART that protects against targeted selection and the bias it can introduce (9). Details can be found elsewhere, but the key innovation in the BCF model is to treat the predicted outcome as a sum of a treatment effect (*τ*) plus the effect of other factors (*μ*). In our context, the other outcome effect (*μ*) arises from other treatments and patient natural history, such as the development of contracture, bony remodeling, neuromaturation, and growth. Both *τ* and *μ* are assumed to depend on a set of chosen covariates and the propensity score. For the BCF estimate we used the bcf function and all default settings except for `ntree_moderate = 200` and `base_moderate = 0.95`. These were increased from their default values (50 and 0.25, respectively) since there is known to be substantial outcome heterogeneity across observations. We used 1000 burn-in MCMC iterations and 1000 MCMC iterations after burn-in.

#### Why use Three Models?

There is an extensive literature describing each of these models and their use. Of note for this study is the work of Hill, who demonstrated the principles by which BART-based models (e.g., VT and BCF) achieve accurate causal predictions (29). This was followed up by the work of Dorie, who compared a large number of state-of-the-art causal inference methods on a large set of challenging datasets (30). Dorie’s study showed that BART-based methods, including BCF, performed exceptionally well and provided more accurate and precise treatment predictions than other causal inference methods. The three methods described vary in approach – though they are not completely independent of one another. Each method also comes with certain assumptions and limitations. For example, the direct matching approach is the most easily understood, and most closely mirrors an RCT, but we can only estimate the treatment effect for observations for which we can identify a matching control. This may result in an effect estimate based on a small or potentially non-representative sample. In contrast, both the VT and BCF models can estimate a treatment effect on every treated observation. However, understanding the mechanism of estimation for the VT and BCF approaches requires significant statistical and algorithmic knowledge, and is harder to understand for clinicians and patients.

### 2.4 Analysis

#### Sample considerations

The DM model produces a set of one-to-one matched treated and control observations (*matched subset*). A limitation of direct matching is that not every treated observation will have a matching control observation. The exclusion of treated limbs creates a risk of bias in the treatment effect estimate. For example, consider a hypothetical situation where more severely affected individuals benefit the most from a treatment but cannot be closely matched to untreated observations because all such severely affected individuals underwent treatment. We look for possible bias from this scenario by estimating a treatment effect for both the *matched subset* and *all treated* observations in the VT and BCF models. While the VT and BCF models can estimate effects for all observations, uncertainty in the regions of poor overlap tends to be large (29,30).

#### Bootstrap bounds

For each model × treatment × outcome combination, the mean and 95% confidence interval for the average treatment effects were derived from 1000 bootstrap replicates sampled from the relevant sets of observations (*matched subset* or *all treated*). Our observations are limbs, so by using bootstrap estimates we avoid making assumptions about the strength of correlation between observations.

## 3 Results

Relevant summary data are provided in this section. In support of transparency and thoroughness, a detailed report for each of the 13 treatments is available as electronic addendum to this manuscript.

### 3.1 Clinical profile

The data for this analysis was from limbs of patients seen for clinical evaluation in our gait analysis laboratory between 2003 and 2020 (inclusive). The entire dataset consisted of 2851 limbs from 933 individuals. After excluding observations with missing covariates or FAQt values we were left with 2502 limbs from 837 individuals [Table 4]. There was no noticeable pattern to the missing data. The main culprits were missing survey data (FAQt, N_miss_ = 224) due to typical non-response rates and maximum passive ankle dorsiflexion with the knee extended (ANK_DORS_0, N_miss_ = 144) due to severe knee flexion contractures. No other variable was missing for more than 24 limbs. Note that the final number of observations for each treatment will be smaller than 2502, and will vary slightly between treatments, due to missing treatment-specific outcome data.

**Table 4.** Participant (limb) characteristics.

| Covariate | Value |
| --- | --- |
| Age in Years (mean (SD)) | 9.4 (3.7) |
| Sex (N Male (%)) | 1453 (58.1) |
| Follow Up Years (mean (SD)) | 1.5 (0.4) |
| Topographic Classification (N (%)) |  |
| *Hemiplegia* | 202 (8.1) |
| *Diplegia* | 1634 (65.3) |
| *Triplegia* | 416 (16.6) |
| *Quadriplegia* | 250 (10.0) |
| GMFCS Level (N (%)) |  |
| *I* | 584 (23.3) |
| *II* | 844 (33.7) |
| *III* | 552 (22.1) |
| *IV* | 19 (0.8) |
| *Missing* | 503 (20.1) |

### 3.2 Treatment Effects

In general, effects were largest at the anatomy and physiology level (borderline medium effect), and decreased to borderline small or none as we moved along the causal chain through gait parameter, overall gait pattern, and function [Figure 1].

**Figure 1.**
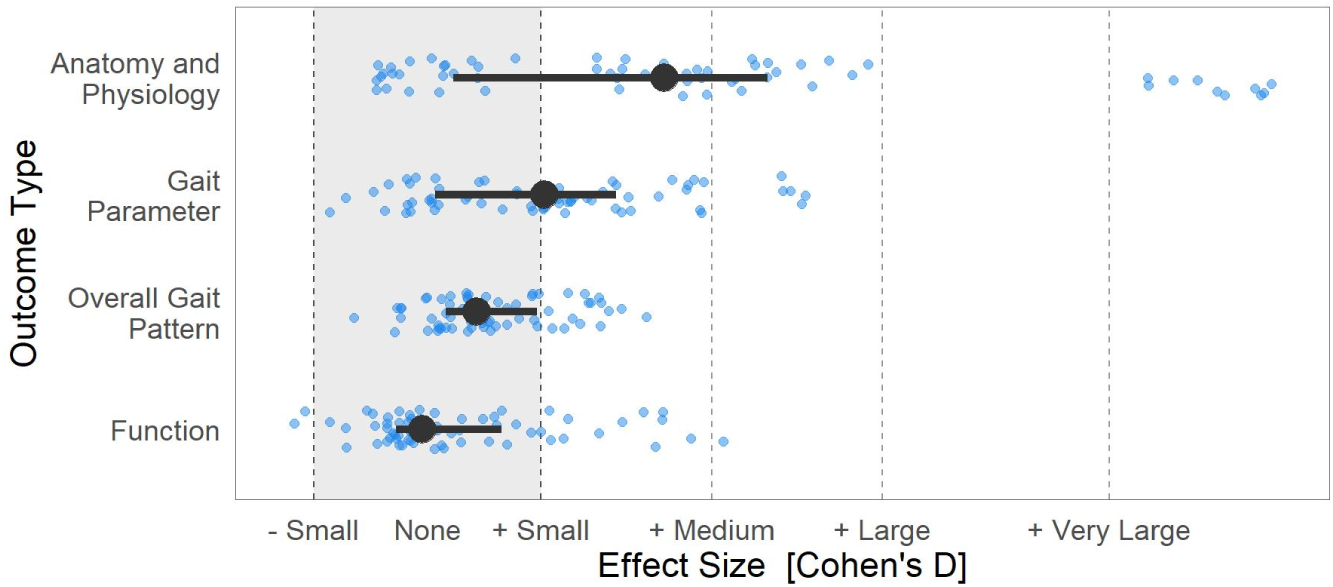
Median and interquartile range of ATT across all treatments, models, and outcomes (large circles and horizontal lines) and individual model estimates across 5 models per outcome (small circles). We use conventional values for effect size thresholds (small ≥ .2, medium ≥ .5, large ≥ .8, very large ≥ 1.2). The very large effects are from Neural Rhizotomy (mean spasticity) and Femoral Derotation Osteotomy (femoral anteversion).

The ATT for each of the 13 treatments at each of the four levels show good consistency across models [Figure 2].

**Figure 2.**
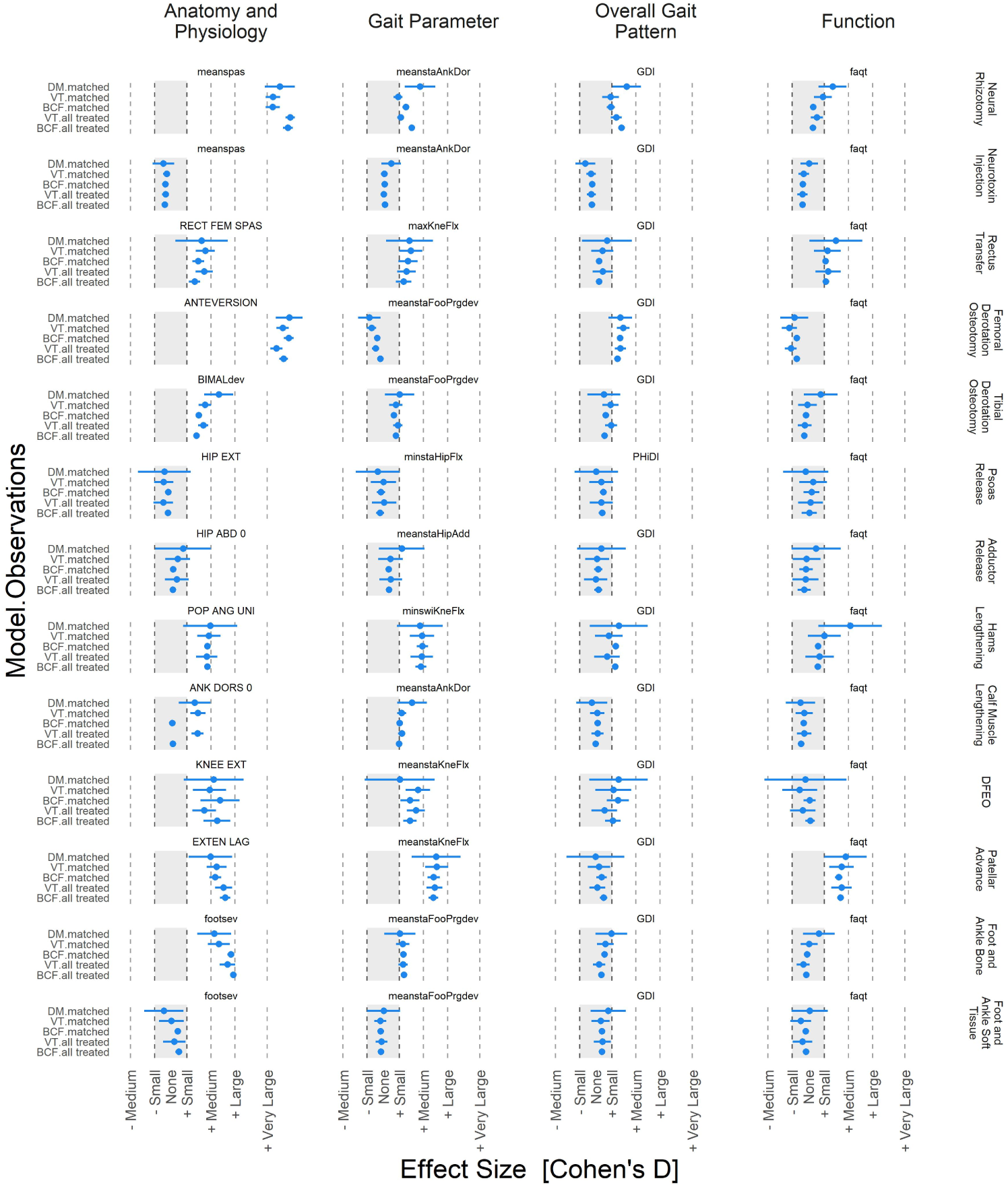
Effect sizes (mean and 95% CI) for 13 treatments, four outcome levels, three models, and two sets of observations. There is good consistency between models and between samples (matched subset and all treated) within a given model (VT or BCF).

There were a few exceptions to this, such as change in mean stance ankle dorsiflexion after Neural Rhizotomy or change in passive ankle dorsiflexion after calf muscle lengthening. However, in these cases the differences were modest, varying within the limits of a single effect category. There were no disagreements among any models, treatments, or outcomes in terms of the sign of the effect. Effect sizes for individual outcomes ranged from very large (e.g., spasticity reduction following selective dorsal rhizotomy) to no effect (many examples). There were no negative ATTs at for any treatment at any level of outcome.

### 3.3 Model performance

To assess the quality of the ATT estimates, we need to examine the performance of the DM, VT, and BCF models.

#### Direct Matching

Matches were obtained for most limbs across all 13 treatments, (mean = 74%, range = 46% -95%) of treated limbs [Table 5].

**Table 5.** Numbers of Treated and Matched Observations.

| **Treatment** | **# Treated** | **# Matched** | **% Matched** |
| --- | --- | --- | --- |
| Neural Rhizotomy | 435 | 200 | 46% |
| Neurotoxin Injection | 522 | 478 | 92% |
| Rectus Transfer | 91 | 79 | 87% |
| Femoral Derotation Osteotomy | 440 | 302 | 69% |
| Tibial Derotation Osteotomy | 316 | 190 | 60% |
| Psoas Release | 96 | 90 | 94% |
| Adductor Release | 78 | 69 | 88% |
| Hams Lengthening | 86 | 76 | 88% |
| Calf Muscle Lengthening | 318 | 210 | 66% |
| DFEO<sup>†</sup> | 75 | 41 | 55% |
| Patellar Advance | 145 | 93 | 64% |
| Foot and Ankle Bone | 336 | 189 | 56% |
| Foot and Ankle Soft Tissue | 147 | 137 | 93% |
| **Mean** | **237** | **166** | **74%** |

<sup>†</sup> *DFEO = Distal Femoral Extension Osteotomy*

The imposed criteria resulted in excellent matching of the physical exam, kinematic, and propensity score covariates, reducing standardized mean differences (SMD) between two- and sixfold [Table 6]. Note that the SMD for factor variables was computed following Yang and Dalton (31).

**Table 6.** Covariate Balance.

| Treatment | Kinematics |  | Propensity |  | Physical Examination |  |
| --- | --- | --- | --- | --- | --- | --- |
|  | Raw | Matched | Raw | Matched | Raw | Matched |
| Neural Rhizotomy | 0.22 (0.23) | 0.08 (0.05) | 0.60 (0.69) | 0.11 (0.11) | 0.30 (0.30) | 0.09 (0.07) |
| Neurotoxin Injection | 0.08 (0.06) | 0.05 (0.03) | 0.23 (0.27) | 0.05 (0.06) | 0.11 (0.11) | 0.06 (0.05) |
| Rectus Transfer | 0.25 (0.20) | 0.10 (0.08) | 1.09 (0.74) | 0.13 (0.10) | 0.35 (0.34) | 0.12 (0.10) |
| Femoral Derotation Osteotomy | 0.14 (0.13) | 0.07 (0.05) | 0.56 (0.50) | 0.11 (0.08) | 0.20 (0.22) | 0.08 (0.06) |
| Tibial Derotation Osteotomy | 0.14 (0.11) | 0.07 (0.06) | 0.69 (0.41) | 0.10 (0.12) | 0.19 (0.21) | 0.08 (0.07) |
| Psoas Release | 0.23 (0.17) | 0.14 (0.10) | 0.99 (0.64) | 0.14 (0.07) | 0.33 (0.31) | 0.15 (0.11) |
| Adductor Release | 0.34 (0.24) | 0.10 (0.07) | 1.05 (0.69) | 0.12 (0.08) | 0.41 (0.34) | 0.12 (0.10) |
| Hams Lengthening | 0.28 (0.21) | 0.16 (0.13) | 0.89 (0.60) | 0.15 (0.09) | 0.33 (0.30) | 0.16 (0.13) |
| Calf Muscle Lengthening | 0.12 (0.08) | 0.08 (0.06) | 0.55 (0.48) | 0.14 (0.10) | 0.18 (0.21) | 0.09 (0.07) |
| DFEO | 0.33 (0.29) | 0.18 (0.13) | 1.40 (1.33) | 0.16 (0.19) | 0.45 (0.52) | 0.19 (0.16) |
| Patellar Advance | 0.27 (0.24) | 0.11 (0.08) | 0.97 (0.78) | 0.13 (0.10) | 0.36 (0.35) | 0.13 (0.10) |
| Foot and Ankle Bone | 0.16 (0.17) | 0.12 (0.09) | 0.69 (0.41) | 0.13 (0.07) | 0.21 (0.25) | 0.11 (0.09) |
| Foot and Ankle Soft Tissue | 0.14 (0.10) | 0.08 (0.07) | 0.63 (0.40) | 0.17 (0.12) | 0.20 (0.22) | 0.11 (0.09) |
| **Mean** | 0.21 | 0.10 | 0.79 | 0.13 | 0.28 | 0.11 |

*All values mean (sd)*

An important result from the DM model is the matching of prior and interval treatment. As noted above, the multi-level nature of surgery in CP means that most patients receive several simultaneous surgeries, many of which can be assumed to have treatment effects across all four levels of outcome [Table 7]. The difference in rates of mean (sd) prior treatment was 3% (3%), and the difference in rates of interval treatment was 3% (3%).

**Table 7.** Balance of Prior and Interval Treatment.

| **Interval Treatment** | **Difference in Prior Treatment Rate (Treated – Control)** |  |  |  |  |  |  |  |  |  |  |  |  |
| --- | --- | --- | --- | --- | --- | --- | --- | --- | --- | --- | --- | --- | --- |
|  | Neural Rhizotomy | Neurotoxin Injection | Rectus Transfer | Femoral Derotation Osteotomy | Tibial Derotation Osteotomy | Psoas Release | Adductor Release | Hams Length. | Calf Muscle Length. | DFEO | Patella Advance | Foot Ankle Bone | Foot Ankle Soft Tiss. |
| Neural Rhizotomy | -10% | -4% | 0% | -4% | -3% | -1% | -1% | -4% | 3% | -1% | -1% | -1% | 3% |
| Neurotoxin Injection | 1% | 5% | 0% | -1% | 1% | 1% | 0% | -3% | -2% | -1% | -1% | 5% | 2% |
| Rectus Transfer | -1% | -9% | -1% | 1% | 1% | 5% | 0% | 5% | 6% | -4% | 0% | -9% | 1% |
| Femoral Derotation Osteotomy | -7% | -3% | 1% | -2% | -2% | -1% | -1% | -1% | -1% | 2% | -1% | -2% | -1% |
| Tibial Derotation Osteotomy | -5% | 0% | -1% | 1% | -2% | 0% | -1% | -1% | -2% | 0% | 2% | -1% | -2% |
| Psoas Release | -6% | 2% | -9% | 1% | 1% | -2% | -8% | -9% | 0% | -9% | -3% | -2% | 11% |
| Adductor Release | 12% | -6% | -4% | 4% | -10% | -1% | -12% | -1% | -3% | -9% | -3% | -3% | -3% |
| Hams Lengthening | -7% | 0% | 4% | 8% | 13% | -4% | 1% | 1% | 8% | 0% | 0% | -3% | -1% |
| Calf Muscle Lengthening | -9% | 3% | 5% | -2% | 0% | 2% | 0% | 0% | -2% | 0% | -1% | 0% | 0% |
| DFEO | 0% | -2% | 2% | 17% | 0% | 2% | 0% | -5% | 0% | 20% | 0% | -2% | 15% |
| Patellar Advance | 8% | -4% | -2% | 3% | -3% | 9% | -2% | 8% | 9% | -4% | 2% | 8% | -5% |
| Foot and Ankle Bone | 5% | 1% | 4% | 3% | 1% | 2% | 0% | 3% | 4% | -3% | 1% | 2% | 2% |
| Foot and Ankle Soft Tissue | -8% | -2% | -1% | 3% | 4% | 3% | 7% | 5% | 3% | -4% | -1% | -2% | -1% |

| **Interval Treatment** | **Difference in Interval Treatment Rate (Treated – Control)** |  |  |  |  |  |  |  |  |  |  |  |  |
| --- | --- | --- | --- | --- | --- | --- | --- | --- | --- | --- | --- | --- | --- |
|  | Neural Rhizotomy | Neurotoxin Injection | Rectus Transfer | Femoral Derotation Osteotomy | Tibial Derotation Osteotomy | Psoas Release | Adductor Release | Hams Length. | Calf Muscle Length. | DFEO | Patellar Advance | Foot Ankle Bone | Foot Ankle Soft Tiss. |
| Neural Rhizotomy | 100% | -2% | -2% | -2% | -2% | -2% | -1% | -1% | -2% | -2% | -2% | -2% | -2% |
| Neurotoxin Injection | -1% | 100% | 1% | 7% | 1% | 0% | 0% | 0% | 2% | 0% | 1% | 1% | 0% |
| Rectus Transfer | -3% | 0% | 100% | 3% | -3% | 4% | 3% | 3% | 5% | 4% | 4% | 4% | 4% |
| Femoral Derotation Osteotomy | -1% | 13% | 5% | 100% | 1% | 4% | 5% | 4% | 1% | 2% | 5% | 1% | -1% |
| Tibial Derotation Osteotomy | -1% | -5% | 2% | 1% | 100% | 4% | 6% | 3% | 1% | -2% | 4% | 3% | 1% |
| Psoas Release | -2% | -7% | 19% | 2% | 3% | 100% | 2% | -1% | 2% | 2% | 2% | 2% | 13% |
| Adductor Release | -3% | 12% | 9% | 4% | 4% | 4% | 100% | 3% | 0% | 4% | 4% | 4% | -1% |
| Hams Lengthening | -3% | -4% | 16% | 3% | 0% | -3% | 3% | 100% | 3% | -3% | 3% | -1% | -1% |
| Calf Muscle Lengthening | -1% | 0% | -1% | 1% | 1% | 1% | -1% | 1% | 100% | -1% | -1% | 1% | 1% |
| DFEO | -5% | 7% | 2% | 12% | 5% | 5% | 2% | -2% | 2% | 100% | 0% | 0% | -10% |
| Patellar Advance | -2% | 6% | 2% | 15% | 12% | 4% | 3% | 1% | 0% | 0% | 100% | 3% | 4% |
| Foot and Ankle Bone | -1% | -6% | 1% | 1% | 2% | 1% | -2% | 1% | 1% | 1% | 1% | 100% | 2% |
| Foot and Ankle Soft Tissue | -1% | -2% | 11% | -1% | 2% | 7% | 4% | -2% | 1% | -1% | 1% | 1% | 100% |

Effects of Common Treatments in CP 1

#### BART Virtual Twins and Bayesian Causal Forests

The key mechanism by which the BART-based models (VT and BCF) provide unbiased ATT estimates is their ability to model a complex outcome response surface as a function of covariates (29). Model accuracy is a surrogate for how well the VT and BCF models fit the response surface. For both the VT and BCF models we examined the predictive accuracy for the selected outcomes [Table 8]. Detailed VT and BCF results are available in the electronic addenda.

**Table 8.** Performance of BART and BCF.

| Outcome | Accuracy (RMSE) |  |
| --- | --- | --- |
|  | BART | BCF |
| ANK_DORS_0 | 6.1 | 6.4 |
| ANTEVERSION | 7.5 | 9.4 |
| BIMALdev | 5.2 | 5.8 |
| EXTEN_LAG | 5.0 | 6.3 |
| faqt (dimensionless) | 11.1 | 11.4 |
| footsev (dimensionless) | 0.32 | 0.32 |
| GDI (dimensionless) | 6.5 | 7.0 |
| HIP_ABD_0 | 5.6 | 6.0 |
| HIP_EXT | 7.9 | 8.2 |
| KNEE_EXT | 3.3 | 3.9 |
| maxKneFlx | 5.2 | 5.7 |
| meanspas | 0.17 | 0.23 |
| meanstaAnkDor | 3.5 | 4.5 |
| meanstaFooPrgdev | 5.8 | 6.7 |
| meanstaHipAdd | 3.9 | 3.9 |
| meanstaKneFlx | 5.6 | 6.4 |
| minstaHipFlx | 5.6 | 5.9 |
| minswiKneFlx | 6.1 | 6.7 |
| PHiDI (dimensionless) | 6.9 | 7.3 |
| POP_ANG_UNI | 8.5 | 9.3 |
| RECT_FEM_SPAS (dimensionless) | 0.27 | 0.36 |

*All units degrees, except where otherwise noted*
*See Appendix 1 for covariate names*

## 4 Discussion

The ATT of 13 common treatments in children and young adults with CP were found to be generally small to medium at the anatomy and physiology level (median [IQR] = 0.42 [.05, .60]) and became smaller along the causal chain through gait parameters (0.21 [.01, .33]), overall gait pattern (0.09 [.03, .19]), and function (−0.01 [-.06, .13]).

### 4.1 Are the models accurate?

The fundamental problem with causal inference is that we can never simultaneously observe an individual under the actual and counterfactual treatment. As a result, we rely on theory and indirect evidence to provide support for the validity our estimates. We implemented modern, widely used, extensively validated approaches. Nevertheless, it is worth looking for indirect evidence for the accuracy of the results. There were two treatments that had large and very large ATTs: selective dorsal rhizotomy (reduction in mean spasticity, Cohen’s d ∼1.5) and femoral derotation osteotomy (reduction in anteversion, Cohen’s d ∼1.5). These ATTs are consistent with what has been found in observational studies as well as what can be reasonably surmised from the nature of the surgeries (sectioning nerve rootlets, large rotation of bone). In contrast, the models found no effect of neurotoxin injections, which is consistent with the very small or absent effects reported at 4-6 weeks post-injection, likely to be even smaller at a one year follow-up (32).

#### Effects at the anatomy and physiology level

Overall, positive effects (including borderline effects) at the anatomy and physiology level were observed in 9 of 13 treatments. Since the anatomy and physiology are being directly manipulated by the treatment, it is not surprising that we observe the largest ATTs at this level. Notable exceptions were neurotoxin injections, lengthening of the psoas or adductors, and soft tissue surgery at the foot and ankle. Foot and ankle soft tissue procedures (e.g., tendon transfers) are primarily prescribed for the correction of dynamic swing-phase foot deformity, so the lack of impact on static, weight-bearing, foot deformity measures is not surprising. Regarding psoas lengthening, previous analyses from our and other institutions have suggested this surgery is only marginally effective (33,34). The adductor data are taken at face value, given that adductor lengthening is often prescribed to correct hip dislocation or subluxation, and there is little research into the effect of this procedure on gait related outcomes.

#### Effects at the gait parameter level

Overall, positive effects at the gait parameter level were observed in 8 of 13 treatments (including three borderline effects). Medium effects were observed for patellar advancement (mean stance knee flexion), followed by borderline medium effects from hamstrings lengthening (minimum swing-phase knee flexion). Small and borderline effects were observed for rhizotomy, rectus transfer, tibial derotation osteotomy, calf muscle lengthening, distal femoral extension osteotomy, and foot and ankle bony surgery. A notable result was the failure of a femoral derotation osteotomy to improve foot progression deviation. This was mostly due to a substantial number of limbs being “overcorrected” (i.e., exhibiting a post-treatment deformity in the opposite direction of the pre-treatment deformity). The risk for overcorrection of limbs has been previously noted, and seems to arise due to an over-reliance on static measures of anteversion in the absence of dynamic signs of internal rotation (35,36).

#### Effects at the overall gait pattern level

Positive effects, including borderline effects, at the overall gait pattern level were observed in 5 of 13 treatments: rhizotomy, femoral derotation osteotomy, tibial derotation osteotomy, distal femoral extension osteotomy, and hamstrings lengthening. The first two are unsurprising, given their large to very large effect at the anatomy and physiology level. The positive hamstrings results were unexpected. Historically, our institution has been conservative in the prescription of hamstrings lengthening surgery. Evidence of this can be seen in Arnold’s study of hamstring lengthening outcomes, where patients from our center comprised the control group of children meeting criteria for hamstrings lengthening but not receiving the treatment (37). One possible explanation of the positive hamstrings result is that, for at least the last 24 years, our center has considered explicit muscle length and lengthening rate thresholds when evaluating candidacy for hamstrings lengthening (38). While muscle length data cannot identify short hamstrings (rule in for surgery), they can identify hamstrings that attain adequate length during gait (rule out for surgery). Using this guidance prevents over-lengthening of non-contracted hamstrings. Importantly, in previous analyses of both psoas and calf muscle lengthening, the largest impact on outcomes arose from ruling out patients who were not good candidates for the surgery (33,39). It seems reasonable to assume the same is true for the hamstrings.

#### Effects at the function level

Overall, positive effects at the function level were observed in 3 of 13 treatments, including two treatments with borderline effects. Patellar tendon advancement had a clear small effect, while rectus femoris transfer and hamstrings lengthening had borderline small effects. A femoral derotation osteotomy had a borderline negative effect – the closest any treatment came to having a negative ATT. While this was effect was not significant, it was surprising, and merits further investigation, given that femoral derotation osteotomy was the most common surgery in this dataset (22% of limbs).

### 4.2 Model performance

#### Matching

The direct matching algorithm worked exceptionally well, considering the complexity of matching physical examination, gait, and treatment covariates. The model was hand tuned to produce a balance between closeness of matching and number of matched observations. We prioritized matching of treatments, due to their importance in outcomes. We were able to achieve excellent results, matching both prior and interval treatments within 3%, on average. We also took care to maintain closely balanced baseline gait kinematic patterns, given the gait-centric nature of this study (mean SMD = 0.21). This was largely achieved with the distance matching, though occasional moment matching parameters were required (e.g., to balance mean stance foot progression for calf muscle lengthening). We were able to match a majority (74%) of treated limbs. Concerns about bias due to omitted observations was addressed by including models that estimated the ATT for all treated limbs, not just the matched subset. Generally, we found no meaningful bias in the matching model.

It is worth reiterating that all propensity models and outcome prediction models used the same set of covariates. This was done in part for simplicity and uniformity of approach, and in part because it reflects the way in which treatment decisions are made in a “*holistic*” manner. Reviewing patient data consists of the iterative identification and assessment of problems based on different domains of the patient profile. It is not possible to identify which factors influence which treatments. Additionally, the use of a uniform set of covariates greatly simplifies external efforts to refute or replicate the findings presented here.

### 4.3 Limitations

With any retrospective study there is a possibility of selection bias. We defend against this with models that can generate ATT estimates on either all, or a subset of treated observations. We did not see meaningful differences between these observations. Another possible source selection bias arises from only evaluating individuals seen for clinical gait analysis and returning for post-treatment evaluation. It is possible that these individuals have a different outcome from either patients not seen for gait analysis at all, or those not seen for follow-up evaluation. Reasonable arguments can be made that these omitted patients could fare better or worse than the sample we analyzed. In either case, it seems unlikely that such a bias would be large.

We used a comprehensive set of covariates, derived from extensive clinical experience and understanding of the underlying condition and mechanisms of treatment effect. We are limited, however, by the measures we routinely obtain for our patients. For example, we do not routinely and objectively measure certain potentially important factors, such as cognitive ability or socioeconomic status. There are logical and scientifically valid reasons to believe that these factors could play a meaningful role in treatment outcome. That said, we suspect that such factors are nearly randomly distributed among the treated and control observations we analyzed, and therefore would not introduce significant bias into our ATT estimates. This issue should be addressed in future work.

We chose a coarse definition of treatments, and therefore cannot examine the importance of varying techniques within a treatment category. In cases where we have studied technique differences within our center and across centers – such as rhizotomy at the conus medullaris versus the cauda equina or proximal versus distal femoral derotation osteotomy – we have not found meaningful differences on outcome (40,41). Our definition of treatments focused on individual procedures, even though multi-level treatment is common. We suspect that there may be an additive effect in SEMLS, although noncomplementary combinations of treatments could exist as well. Applying the methods described here to SEMLS (two or more treatments on a given limb) and using no treatment as a control, we observed a median effect across all models and samples of 0.36 for GDI (between small and medium) and 0.02 for FAQt (no effect).

Finally, the results presented here represent the short-term outcome at approximately one-year follow-up. Long-term and immediate impacts merit further investigation.

### 4.4 Future Work

The replication crisis in science is real. Thus, generalizability to other centers needs to be tested. We intentionally chose commonly measured variables and broad treatment categories in service to this goal. We also used out-of-the-box R packages that are open-source, easy-to-use, fast, stable, and well documented. Finally, to aid other researchers, the electronic addenda to this manuscript include extensive and detailed descriptions of the modeling parameter choices and intermediate results. Detailed code and data can be provided upon reasonable request.

Communication to clinicians and patients remains a critical challenge for machine learning methods in medicine. It is important that the consumers of this information understand the strengths and weaknesses of the approaches so that they can make informed decisions based on the results. The DM approach is the easiest to understand and most intuitive of the three models. It was therefore included, even though it has the widest uncertainty bounds is at greatest risk for bias due to omitted observations.

This study examined average effects for broad treatment categories. Finding factors that explain heterogeneous treatment effects (HTEs) is an obvious next step (42). Beyond HTEs lies the last step in the most holy grail of personalized medicine – predicting individual treatment effects (ITEs). All three levels of analysis have strengths and weaknesses. While the ATT analysis is likely to be the most accurate and generalizable, it provides the least specific guidance for an individual patient. In contrast, ITEs could be valuable to the clinician and patient, but are likely to have extremely wide uncertainty bounds. Despite its limitations, the ATT level of forecasting is a significant improvement over the current norm in treatment of children and young adults with CP, which generally does not include any explicit guidance, and instead relies mostly on clinician experience, intuition, and local treatment culture.

## Supporting information

Detailed Results for Each of the 13 Treatments

## Data Availability

The datasets generated during and/or analysed during the current study are not publicly available due to patient confidentiality, but are available from the corresponding author on reasonable request.

## Appendix 1. Variable names

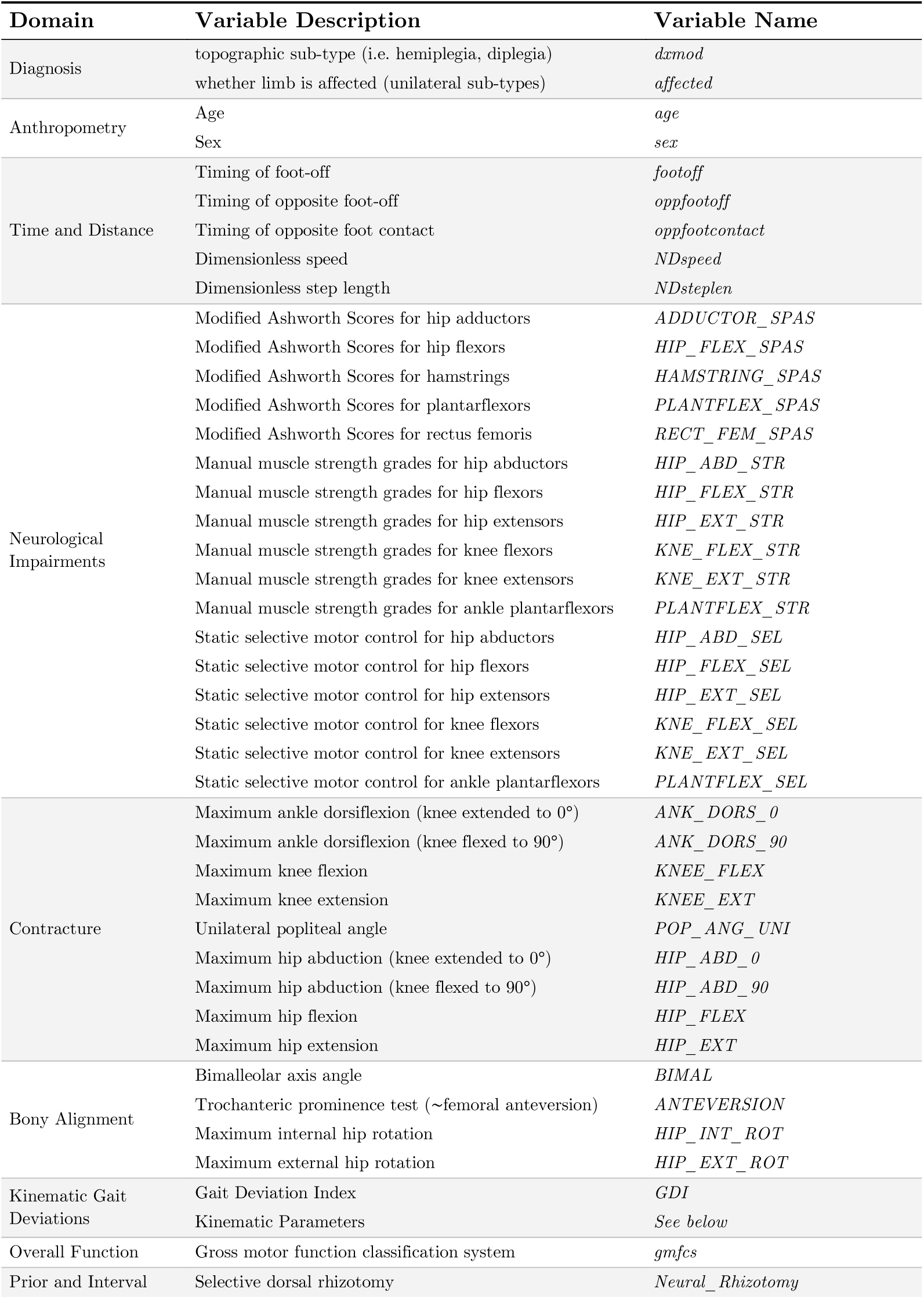
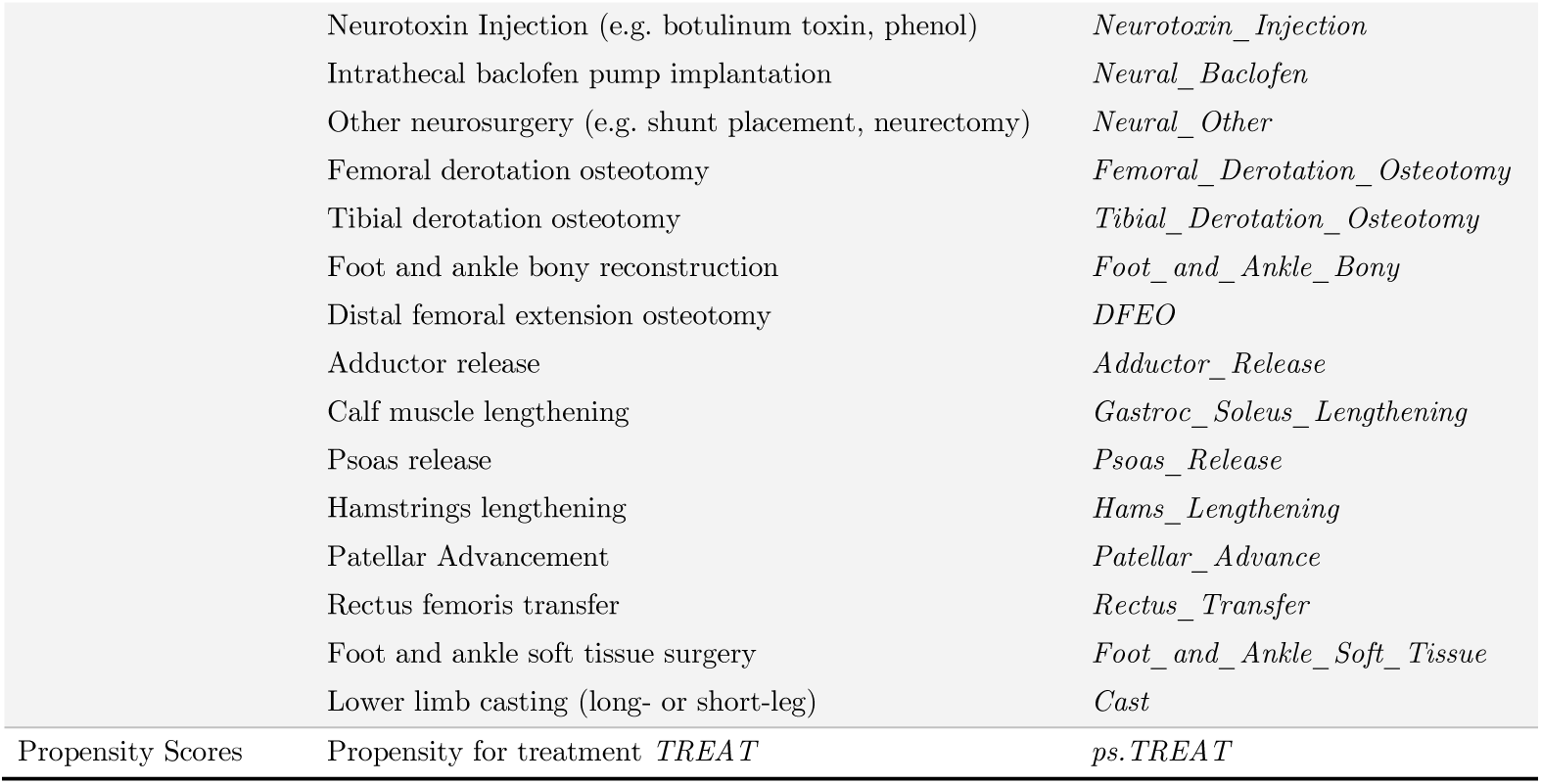

### Naming of kinematic parameters

#### Kinematic sign conventions and abbreviations

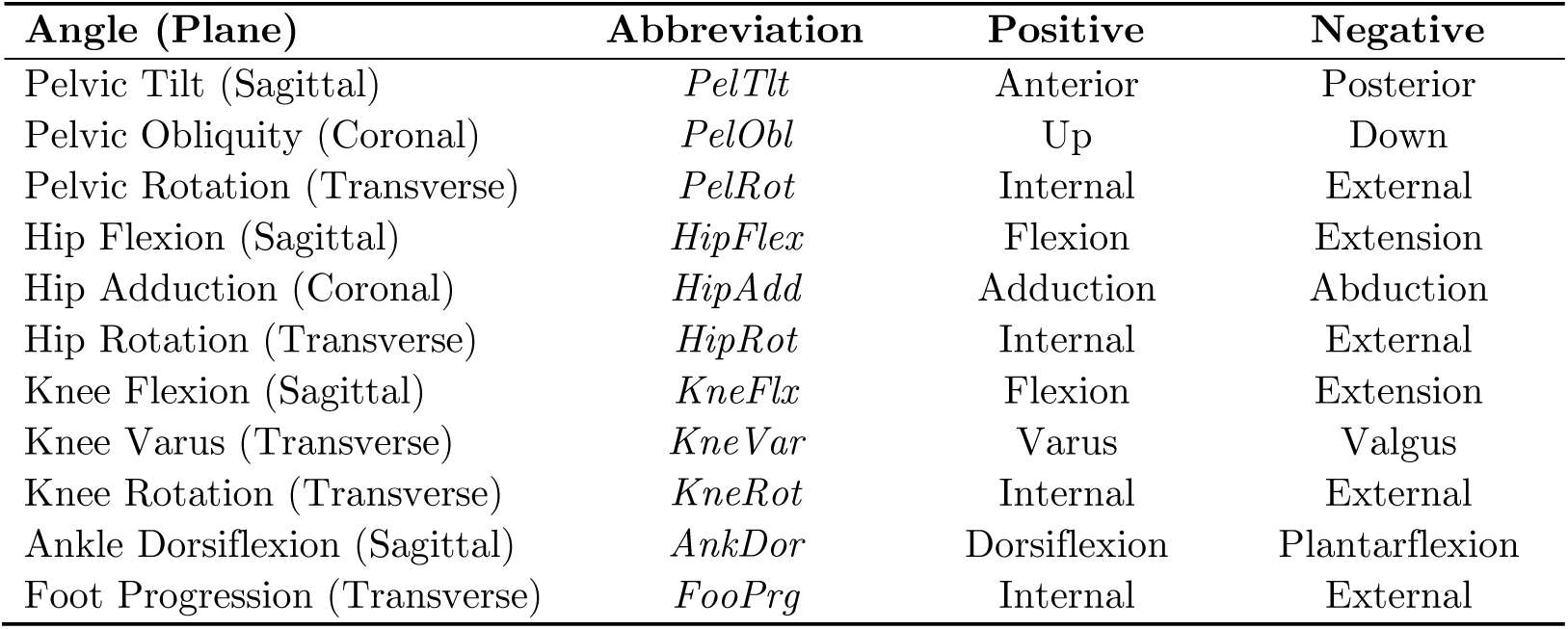

#### Kinematic parameters are named by level, plane, and measure

- ***Level*** can be pelvis, hip, knee, ankle/foot
- ***Plane*** can be sagittal, coronal, or transverse
- ***Measure***
  - Angle (maximum, minimum, mean, initial contact value, foot-off value)
  - Timing of an angle (time of maximum, time of minimum),
  - Defined over stance-phase, swing-phase, or the entire gait cycle

#### Below are some examples

- ***Level*** = pelvis, ***Plane*** = coronal, ***Measure*** = minimum in stance minimum stance-phase pelvic obliquity (*minstaPelObl*).
- ***Level*** = knee, ***Plane*** = sagittal, ***Measure*** = timing of max in swing timing of maximum swing-phase knee flexion (*t_maxswiKneFlx*)
- ***Level*** = hip, ***Plane*** = transverse, ***Measure*** = value at initial contact initial contact hip rotation (*icHipRot*)
- ***Level*** = foot, ***Plane*** = transverse, ***Measure*** = mean over gait cycle mean foot progression (*meanFooPrg*)

Note that there are no measures for ***Level*** = ankle and foot and ***Plane*** = coronal.

## Notes

### Competing Interest Statement

The authors have declared no competing interest.

### Funding Statement

Funding provided by the generous donors of Gillette Children's Specialty Healthcare

### Author Declarations

IRB of University of Minnesota gave ethical approval for this work

### Summary of Updates

Added electronica addenda

## References

1. Yeargin-Allsopp M, Van Naarden Braun K, Doernberg NS, Benedict RE, Kirby RS, Durkin MS. Prevalence of cerebral palsy in 8-year-old children in three areas of the United States in 2002: a multisite collaboration. Pediatrics. 2008 Mar;121(3):547–54.

2. Kirby RS, Wingate MS, Van Naarden Braun K, Doernberg NS, Arneson CL, Benedict RE, et al. Prevalence and functioning of children with cerebral palsy in four areas of the United States in 2006: a report from the Autism and Developmental Disabilities Monitoring Network. Res Dev Disabil. 2011 Apr;32(2):462–9.

3. Christensen D, Van Naarden Braun K, Doernberg NS, Maenner MJ, Arneson CL, Durkin MS, et al. Prevalence of cerebral palsy, co-occurring autism spectrum disorders, and motor functioning - Autism and Developmental Disabilities Monitoring Network, USA, 2008. Dev Med Child Neurol. 2014 Jan;56(1):59–65.

4. Kancherla V, Amendah DD, Grosse SD, Yeargin-Allsopp M, Van Naarden Braun K. Medical expenditures attributable to cerebral palsy and intellectual disability among Medicaid-enrolled children. Res Dev Disabil. 2012 Jun;33(3):832–40.

5. Data and Statistics for Cerebral Palsy | CDC [Internet]. [cited 2021 Sep 9]. Available from: https://www.cdc.gov/ncbddd/cp/data.html

6. Wu YW, Mehravari AS, Numis AL, Gross P. Cerebral palsy research funding from the National Institutes of Health, 2001 to 2013. Dev Med Child Neurol. 2015 Oct;57(10):936–41.

7. Sees JP, Truong WH, Novacheck TF, Miller F, Georgiadis AG. What’s New in the Orthopaedic Treatment of Ambulatory Children With Cerebral Palsy Using Gait Analysis. J Pediatr Orthop. 2020 Jul;40(6):e498–503.

8. Rubin DB. Teaching Statistical Inference for Causal Effects in Experiments and Observational Studies. J Educ Behav Stat. 2004;29(3):343–67.

9. Hahn PR, Murray JS, Carvalho CM. Bayesian Regression Tree Models for Causal Inference: Regularization, Confounding, and Heterogeneous Effects (with Discussion). Bayesian Anal. 2020 Sep;15(3):965–1056.

10. Pearl J. The seven tools of causal inference, with reflections on machine learning. Commun ACM. 2019 Feb 21;62(3):54–60.

11. Yao L, Chu Z, Li S, Li Y, Gao J, Zhang A. A Survey on Causal Inference. 200202770 Cs Stat [Internet]. 2020 Feb 5 [cited 2021 Feb 16]; Available from: http://arxiv.org/abs/2002.02770

12. Schwartz MH, Ries AJ. Rectus femoris transfer in children with cerebral palsy: comparing a propensity score-matched observational study to a randomized controlled trial. Dev Med Child Neurol. 2021;63(2):196–203.

13. Rajagopal A, Kidzinski L, McGlaughlin AS, Hicks JL, Delp SL, Schwartz MH. Estimating the effect size of surgery to improve walking in children with cerebral palsy from retrospective observational clinical data. Sci Rep. 2018 Nov 5;8(1):16344.

14. Holland PW. Statistics and Causal Inference. J Am Stat Assoc. 1986 Dec 1;81(396):945–60.

15. Rubin DB. Estimating Causal Effects of Treatments in Randomized and Nonrandomized Studies. J Educ Psychol [Internet]. 1974 [cited 2021 Aug 31]; Available from: https://dash.harvard.edu/handle/1/3408692

16. Ehrig RM, Taylor WR, Duda GN, Heller MO. A survey of formal methods for determining functional joint axes. J Biomech. 2007;40(10):2150–7.

17. Harris GF, Smith PA. Foot and Ankle Motion Analysis: Clinical Treatment and Technology [Internet]. 0 ed. CRC Press; 2007 [cited 2021 Feb 16]. Available from: https://www.taylorfrancis.com/books/9781420005745

18. Baker R, Finney L, Orr J. A new approach to determine the hip rotation profile from clinical gait analysis data. Hum Mov Sci. 1999 Oct 1;18(5):655–67.

19. Bohannon RW, Smith MB. Interrater reliability of a modified Ashworth scale of muscle spasticity. Phys Ther. 1987 Feb;67(2):206–7.

20. Kendall HO, Kendall FP, Wadsworth GE. Muscles, Testing and Function. Am J Phys Med Rehabil. 1973 Feb;52(1):43.

21. Schwartz MH, Aldahondo N, MacWilliams BA. A Patient-Reported Measure of Locomotor Function Derived from the Functional Assessment Questionnaire. medRxiv. 2021 Jun 16;2021.06.12.21258826.

22. Gorton GE, Stout JL, Bagley AM, Bevans K, Novacheck TF, Tucker CA. Gillette Functional Assessment Questionnaire 22-item skill set: factor and Rasch analyses. Dev Med Child Neurol. 2011;53(3):250–5.

23. Buuren S van, Groothuis-Oudshoorn K. mice: Multivariate Imputation by Chained Equations in R. J Stat Softw. 2011 Dec 12;45(1):1–67.

24. Kapelner A, Bleich J. bartMachine: Machine Learning with Bayesian Additive Regression Trees. J Stat Softw. 2016;70(4):1–40.

25. Austin PC. An Introduction to Propensity Score Methods for Reducing the Effects of Confounding in Observational Studies. Multivar Behav Res. 2011 May 31;46(3):399– 424.

26. Lamberts RP, Burger M, Toit J du, Langerak NG. A Systematic Review of the Effects of Single-Event Multilevel Surgery on Gait Parameters in Children with Spastic Cerebral Palsy. PLOS ONE. 2016 Oct 18;11(10):e0164686.

27. R Core Team. R: A Language and Environment for Statistical Computing [Internet]. Vienna, Austria: R Foundation for Statistical Computing; 2020. Available from: https://www.R-project.org/

28. Zubizarreta JR, Kilcioglu C, Vielma JP. designmatch: Matched Samples that are Balanced and Representative by Design [Internet]. 2018. Available from: https://CRAN.R-project.org/package=designmatch

29. Hill JL. Bayesian Nonparametric Modeling for Causal Inference. J Comput Graph Stat. 2011 Jan 1;20(1):217–40.

30. Dorie V, Hill J, Shalit U, Scott M, Cervone D. Automated versus Do-It-Yourself Methods for Causal Inference: Lessons Learned from a Data Analysis Competition. Stat Sci. 2019 Feb;34(1):43–68.

31. Yang D, Dalton, JE. A Unified Approach to Measuring the Effect Size Between Two Groups Using SAS. In: Proceedings of SAS Global Forum. 2012.

32. Allergan. BOTOX® Treatment in Pediatric Lower Limb Spasticity: Double-blind Study [Internet]. clinicaltrials.gov; 2018 Jul [cited 2021 Sep 13]. Report No.: results/NCT01603628. Available from: https://clinicaltrials.gov/ct2/show/results/NCT01603628

33. Schwartz MH, Rozumalski A, Truong W, Novacheck TF. Predicting the outcome of intramuscular psoas lengthening in children with cerebral palsy using preoperative gait data and the random forest algorithm. Gait Posture. 2013 Apr;37(4):473–9.

34. Sutherland DH, Zilberfarb JL, Kaufman KR, Wyatt MP, Chambers HG. Psoas Release at the Pelvic Brim in Ambulatory Patients with Cerebral Palsy: Operative Technique and Functional Outcome. J Pediatr Orthop. 1997 Oct;17(5):563–70.

35. Schwartz MH, Rozumalski A, Novacheck TF. Femoral derotational osteotomy: Surgical indications and outcomes in children with cerebral palsy. Gait Postur 2014 Feb 1;39(2):778–83.

36. Dreher T, Wolf S, Braatz F, Patikas D, Döderlein L. Internal rotation gait in spastic diplegia--critical considerations for the femoral derotation osteotomy. Gait Posture. 2007 Jun;26(1):25–31.

37. Arnold AS, Liu MQ, Schwartz MH, Ounpuu S, Delp SL. The role of estimating muscle-tendon lengths and velocities of the hamstrings in the evaluation and treatment of crouch gait. Gait Posture. 2006 Apr;23(3):273–81.

38. Schutte LM, Hayden SW, Gage JR. Lengths of hamstrings and psoas muscles during crouch gait: effects of femoral anteversion. J Orthop Res Off Publ Orthop Res Soc. 1997 Jul;15(4):615–21.

39. Rajagopal A, Kidzinski L, McGlaughlin AS, Hicks JL, Delp SL, Schwartz MH. Pre-operative gastrocnemius lengths in gait predict outcomes following gastrocnemius lengthening surgery in children with cerebral palsy. PloS One. 2020;15(6):e0233706.

40. Niklasch M, Boyer ER, Novacheck T, Dreher T, Schwartz M. Proximal versus distal femoral derotation osteotomy in bilateral cerebral palsy. Dev Med Child Neurol. 2018 Oct;60(10):1033–7.

41. Duffy EA, Hornung AL, Chen BP-J, Munger ME, Aldahondo N, Krach LE, et al. Comparing short-term outcomes between conus medullaris and cauda equina surgical techniques of selective dorsal rhizotomy. Dev Med Child Neurol. 2021 Mar;63(3):336– 42.

42. Schwartz MH, Kainz H, Georgiadis AG. Estimating Causal Treatment Effects of Femoral and Tibial Derotational Osteotomies on Foot Progression in Children with Cerebral Palsy. medRxiv. 2021 Mar 8;2021.03.04.21252476.

