## Supplementary material for "Estimating the Efficacy of Common Treatments in Children and Young Adults Diagnosed with Cerebral Palsy Using Three Machine Learning Algorithms": Detailed Results for Each of the 13 Treatments: Foot_and_Ankle_Soft_Tissue.html

#### Matched pairs

|  | Control | Treated | p |
| --- | --- | --- | --- |
| n | 137 | 137 |  |
| AGE (mean (SD)) | 10.60 (2.88) | 10.25 (3.27) | 0.348 |
| GDI (mean (SD)) | 74.26 (9.40) | 73.10 (10.93) | 0.346 |
| NDspeed (mean (SD)) | 0.33 (0.10) | 0.33 (0.08) | 0.974 |
| NDsteplen (mean (SD)) | 0.65 (0.14) | 0.64 (0.13) | 0.498 |
| footoff (mean (SD)) | 0.62 (0.05) | 0.62 (0.04) | 0.704 |
| oppfootoff (mean (SD)) | 0.13 (0.05) | 0.13 (0.04) | 0.975 |
| oppfootcontact (mean (SD)) | 0.49 (0.03) | 0.50 (0.03) | 0.009 |
| ANTEVERSION (mean (SD)) | 39.72 (16.01) | 37.50 (18.25) | 0.287 |
| BIMAL (mean (SD)) | 18.54 (11.37) | 15.12 (13.18) | 0.022 |
| HIP\_INT\_ROT (mean (SD)) | 62.09 (13.05) | 61.20 (14.71) | 0.597 |
| HIP\_EXT\_ROT (mean (SD)) | 32.21 (14.30) | 33.98 (15.17) | 0.322 |
| HIP\_ABD\_0 (mean (SD)) | 32.90 (10.69) | 30.47 (8.89) | 0.042 |
| HIP\_ABD\_90 (mean (SD)) | 64.90 (11.83) | 62.96 (12.18) | 0.181 |
| HIP\_FLEX (mean (SD)) | 129.26 (10.63) | 129.06 (16.92) | 0.905 |
| HIP\_EXT (mean (SD)) | 2.38 (10.73) | 2.44 (11.27) | 0.965 |
| KNEE\_EXT (mean (SD)) | 0.39 (6.09) | 0.03 (6.27) | 0.636 |
| KNEE\_FLEX (mean (SD)) | 134.38 (8.93) | 134.58 (8.63) | 0.847 |
| POP\_ANG\_UNI (mean (SD)) | 54.39 (14.86) | 52.41 (15.32) | 0.277 |
| ANK\_DORS\_0 (mean (SD)) | -3.14 (8.62) | -2.15 (9.98) | 0.379 |
| ANK\_DORS\_90 (mean (SD)) | 7.23 (8.60) | 9.30 (10.53) | 0.076 |
| foAnkDor (mean (SD)) | -4.48 (10.69) | -6.21 (12.93) | 0.228 |
| SEX (mean (SD)) | 0.57 (0.50) | 0.62 (0.49) | 0.391 |
| dx = Cerebral palsy (%) | 137 (100.0) | 137 (100.0) | NA |
| dxmod (%) |  |  | 0.658 |
| Diplegia | 81 ( 59.1) | 76 ( 55.5) |  |
| Hemiplegia | 11 ( 8.0) | 7 ( 5.1) |  |
| Hemiplegia type II | 1 ( 0.7) | 1 ( 0.7) |  |
| Hemiplegia type III | 0 ( 0.0) | 1 ( 0.7) |  |
| Hemiplegia type IV | 3 ( 2.2) | 1 ( 0.7) |  |
| Quadriplegia | 22 ( 16.1) | 27 ( 19.7) |  |
| Triplegia | 19 ( 13.9) | 24 ( 17.5) |  |
| dxside (%) |  |  | 0.465 |
| Asymmetrical L | 27 ( 19.7) | 19 ( 13.9) |  |
| Asymmetrical R | 14 ( 10.2) | 23 ( 16.8) |  |
| Bilateral | 65 ( 47.4) | 66 ( 48.2) |  |
| Left | 11 ( 8.0) | 7 ( 5.1) |  |
| Right | 14 ( 10.2) | 16 ( 11.7) |  |
| Unknown | 6 ( 4.4) | 6 ( 4.4) |  |
| affected (mean (SD)) | 0.89 (0.31) | 0.93 (0.26) | 0.296 |
| gmfcs (%) |  |  | 0.681 |
| 1 | 26 ( 19.0) | 24 ( 17.5) |  |
| 2 | 51 ( 37.2) | 50 ( 36.5) |  |
| 3 | 27 ( 19.7) | 25 ( 18.2) |  |
| 4 | 0 ( 0.0) | 2 ( 1.5) |  |
| Miss | 33 ( 24.1) | 36 ( 26.3) |  |
| HIP\_ABD\_SEL (%) |  |  | 0.448 |
| 0 | 12 ( 8.8) | 10 ( 7.3) |  |
| 1 | 64 ( 46.7) | 54 ( 39.4) |  |
| 2 | 53 ( 38.7) | 60 ( 43.8) |  |
| Miss | 8 ( 5.8) | 13 ( 9.5) |  |
| HIP\_EXT\_SEL (%) |  |  | 0.532 |
| 0 | 9 ( 6.6) | 15 ( 10.9) |  |
| 1 | 53 ( 38.7) | 53 ( 38.7) |  |
| 2 | 64 ( 46.7) | 56 ( 40.9) |  |
| Miss | 11 ( 8.0) | 13 ( 9.5) |  |
| HIP\_FLEX\_SEL (%) |  |  | 0.117 |
| 0 | 2 ( 1.5) | 7 ( 5.1) |  |
| 1 | 73 ( 53.3) | 76 ( 55.5) |  |
| 2 | 55 ( 40.1) | 42 ( 30.7) |  |
| Miss | 7 ( 5.1) | 12 ( 8.8) |  |
| KNEE\_EXT\_SEL (%) |  |  | 0.780 |
| 0 | 11 ( 8.0) | 12 ( 8.8) |  |
| 1 | 62 ( 45.3) | 61 ( 44.5) |  |
| 2 | 57 ( 41.6) | 53 ( 38.7) |  |
| Miss | 7 ( 5.1) | 11 ( 8.0) |  |
| KNEE\_FLEX\_SEL (%) |  |  | 0.045 |
| 0 | 10 ( 7.3) | 21 ( 15.3) |  |
| 1 | 76 ( 55.5) | 59 ( 43.1) |  |
| 2 | 45 ( 32.8) | 45 ( 32.8) |  |
| Miss | 6 ( 4.4) | 12 ( 8.8) |  |
| PLANTFLEX\_SEL (%) |  |  | 0.328 |
| 0 | 30 ( 21.9) | 23 ( 16.8) |  |
| 1 | 82 ( 59.9) | 80 ( 58.4) |  |
| 2 | 16 ( 11.7) | 17 ( 12.4) |  |
| Miss | 9 ( 6.6) | 17 ( 12.4) |  |
| HIP\_ABD\_STR (%) |  |  | 0.371 |
| 2 | 33 ( 24.1) | 31 ( 22.6) |  |
| 3 | 62 ( 45.3) | 70 ( 51.1) |  |
| 4 | 31 ( 22.6) | 22 ( 16.1) |  |
| 5 | 3 ( 2.2) | 1 ( 0.7) |  |
| Miss | 8 ( 5.8) | 13 ( 9.5) |  |
| HIP\_EXT\_STR (%) |  |  | 0.954 |
| 1 | 2 ( 1.5) | 2 ( 1.5) |  |
| 2 | 35 ( 25.5) | 38 ( 27.7) |  |
| 3 | 39 ( 28.5) | 42 ( 30.7) |  |
| 4 | 38 ( 27.7) | 32 ( 23.4) |  |
| 5 | 12 ( 8.8) | 10 ( 7.3) |  |
| Miss | 11 ( 8.0) | 13 ( 9.5) |  |
| HIP\_FLEX\_STR (%) |  |  | 0.671 |
| 3 | 26 ( 19.0) | 25 ( 18.2) |  |
| 4 | 65 ( 47.4) | 60 ( 43.8) |  |
| 5 | 39 ( 28.5) | 40 ( 29.2) |  |
| Miss | 7 ( 5.1) | 12 ( 8.8) |  |
| KNEE\_EXT\_STR (%) |  |  | 0.223 |
| 2 | 2 ( 1.5) | 4 ( 2.9) |  |
| 3 | 49 ( 35.8) | 35 ( 25.5) |  |
| 4 | 18 ( 13.1) | 27 ( 19.7) |  |
| 5 | 61 ( 44.5) | 60 ( 43.8) |  |
| Miss | 7 ( 5.1) | 11 ( 8.0) |  |
| KNEE\_FLEX\_STR (%) |  |  | 0.443 |
| 2 | 6 ( 4.4) | 7 ( 5.1) |  |
| 3 | 40 ( 29.2) | 44 ( 32.1) |  |
| 4 | 69 ( 50.4) | 56 ( 40.9) |  |
| 5 | 16 ( 11.7) | 18 ( 13.1) |  |
| Miss | 6 ( 4.4) | 12 ( 8.8) |  |
| PLANTFLEX\_STR (%) |  |  | 0.666 |
| 0 | 1 ( 0.7) | 1 ( 0.7) |  |
| 1 | 11 ( 8.0) | 9 ( 6.6) |  |
| 2 | 77 ( 56.2) | 71 ( 51.8) |  |
| 3 | 30 ( 21.9) | 32 ( 23.4) |  |
| 4 | 9 ( 6.6) | 7 ( 5.1) |  |
| Miss | 9 ( 6.6) | 17 ( 12.4) |  |
| ADDUCTOR\_SPAS (%) |  |  | 0.320 |
| 1 | 104 ( 75.9) | 91 ( 66.4) |  |
| 2 | 18 ( 13.1) | 25 ( 18.2) |  |
| 3 | 14 ( 10.2) | 18 ( 13.1) |  |
| 4 | 1 ( 0.7) | 3 ( 2.2) |  |
| HAMSTRING\_SPAS (%) |  |  | 0.295 |
| 1 | 107 ( 78.1) | 96 ( 70.1) |  |
| 2 | 24 ( 17.5) | 33 ( 24.1) |  |
| 3 | 5 ( 3.6) | 8 ( 5.8) |  |
| 4 | 1 ( 0.7) | 0 ( 0.0) |  |
| HIP\_FLEX\_SPAS (%) |  |  | 0.029 |
| 1 | 121 ( 88.3) | 106 ( 77.4) |  |
| 2 | 15 ( 10.9) | 25 ( 18.2) |  |
| 3 | 1 ( 0.7) | 6 ( 4.4) |  |
| PLANTFLEX\_SPAS (%) |  |  | 0.467 |
| 1 | 86 ( 62.8) | 81 ( 59.1) |  |
| 2 | 23 ( 16.8) | 27 ( 19.7) |  |
| 3 | 21 ( 15.3) | 20 ( 14.6) |  |
| 4 | 7 ( 5.1) | 6 ( 4.4) |  |
| Miss | 0 ( 0.0) | 3 ( 2.2) |  |
| RECT\_FEM\_SPAS (%) |  |  | 0.028 |
| 1 | 97 ( 70.8) | 78 ( 56.9) |  |
| 2 | 26 ( 19.0) | 34 ( 24.8) |  |
| 3 | 13 ( 9.5) | 17 ( 12.4) |  |
| 4 | 1 ( 0.7) | 8 ( 5.8) |  |
| footsev (mean (SD)) | 0.85 (0.44) | 0.82 (0.39) | 0.593 |
| meanstaFooPrgdev (mean (SD)) | 15.78 (11.26) | 16.83 (13.86) | 0.494 |
| faqt (mean (SD)) | 46.07 (22.60) | 43.18 (23.26) | 0.298 |

#### Unmatched treated vs. Matched treated

This comparison is useful for understanding possible bias from failing to match some treated observations

|  | Unmatched | Matched | p |
| --- | --- | --- | --- |
| n | 10 | 137 |  |
| AGE (mean (SD)) | 9.31 (2.60) | 10.25 (3.27) | 0.375 |
| GDI (mean (SD)) | 66.90 (12.15) | 73.10 (10.93) | 0.088 |
| NDspeed (mean (SD)) | 0.36 (0.08) | 0.33 (0.08) | 0.224 |
| NDsteplen (mean (SD)) | 0.64 (0.15) | 0.64 (0.13) | 0.940 |
| footoff (mean (SD)) | 0.58 (0.06) | 0.62 (0.04) | 0.005 |
| oppfootoff (mean (SD)) | 0.13 (0.05) | 0.13 (0.04) | 0.687 |
| oppfootcontact (mean (SD)) | 0.47 (0.02) | 0.50 (0.03) | 0.001 |
| ANTEVERSION (mean (SD)) | 44.00 (16.80) | 37.50 (18.25) | 0.277 |
| BIMAL (mean (SD)) | 22.00 (15.31) | 15.12 (13.18) | 0.117 |
| HIP\_INT\_ROT (mean (SD)) | 69.00 (14.49) | 61.20 (14.71) | 0.107 |
| HIP\_EXT\_ROT (mean (SD)) | 29.50 (11.65) | 33.98 (15.17) | 0.363 |
| HIP\_ABD\_0 (mean (SD)) | 36.50 (8.83) | 30.47 (8.89) | 0.040 |
| HIP\_ABD\_90 (mean (SD)) | 70.50 (8.32) | 62.96 (12.18) | 0.056 |
| HIP\_FLEX (mean (SD)) | 119.50 (42.13) | 129.06 (16.92) | 0.136 |
| HIP\_EXT (mean (SD)) | 1.50 (12.26) | 2.44 (11.27) | 0.801 |
| KNEE\_EXT (mean (SD)) | 3.00 (7.15) | 0.03 (6.27) | 0.155 |
| KNEE\_FLEX (mean (SD)) | 135.50 (2.84) | 134.58 (8.63) | 0.739 |
| POP\_ANG\_UNI (mean (SD)) | 47.50 (13.39) | 52.41 (15.32) | 0.326 |
| ANK\_DORS\_0 (mean (SD)) | -9.50 (6.43) | -2.15 (9.98) | 0.023 |
| ANK\_DORS\_90 (mean (SD)) | -0.50 (7.25) | 9.30 (10.53) | 0.004 |
| foAnkDor (mean (SD)) | 0.97 (13.88) | -6.21 (12.93) | 0.094 |
| SEX (mean (SD)) | 0.60 (0.52) | 0.62 (0.49) | 0.899 |
| dx = Cerebral palsy (%) | 10 (100.0) | 137 (100.0) | NA |
| dxmod (%) |  |  | <0.001 |
| Diplegia | 2 ( 20.0) | 76 ( 55.5) |  |
| Hemiplegia | 4 ( 40.0) | 7 ( 5.1) |  |
| Hemiplegia type II | 1 ( 10.0) | 1 ( 0.7) |  |
| Hemiplegia type III | 0 ( 0.0) | 1 ( 0.7) |  |
| Hemiplegia type IV | 1 ( 10.0) | 1 ( 0.7) |  |
| Quadriplegia | 0 ( 0.0) | 27 ( 19.7) |  |
| Triplegia | 2 ( 20.0) | 24 ( 17.5) |  |
| dxside (%) |  |  | <0.001 |
| Asymmetrical L | 1 ( 10.0) | 19 ( 13.9) |  |
| Asymmetrical R | 1 ( 10.0) | 23 ( 16.8) |  |
| Bilateral | 2 ( 20.0) | 66 ( 48.2) |  |
| Left | 5 ( 50.0) | 7 ( 5.1) |  |
| Right | 1 ( 10.0) | 16 ( 11.7) |  |
| Unknown | 0 ( 0.0) | 6 ( 4.4) |  |
| affected (mean (SD)) | 0.40 (0.52) | 0.93 (0.26) | <0.001 |
| gmfcs (%) |  |  | 0.102 |
| 1 | 5 ( 50.0) | 24 ( 17.5) |  |
| 2 | 2 ( 20.0) | 50 ( 36.5) |  |
| 3 | 0 ( 0.0) | 25 ( 18.2) |  |
| 4 | 0 ( 0.0) | 2 ( 1.5) |  |
| Miss | 3 ( 30.0) | 36 ( 26.3) |  |
| HIP\_ABD\_SEL (%) |  |  | 0.989 |
| 0 | 1 ( 10.0) | 10 ( 7.3) |  |
| 1 | 4 ( 40.0) | 54 ( 39.4) |  |
| 2 | 4 ( 40.0) | 60 ( 43.8) |  |
| Miss | 1 ( 10.0) | 13 ( 9.5) |  |
| HIP\_EXT\_SEL (%) |  |  | 0.730 |
| 0 | 1 ( 10.0) | 15 ( 10.9) |  |
| 1 | 4 ( 40.0) | 53 ( 38.7) |  |
| 2 | 3 ( 30.0) | 56 ( 40.9) |  |
| Miss | 2 ( 20.0) | 13 ( 9.5) |  |
| HIP\_FLEX\_SEL (%) |  |  | 0.641 |
| 0 | 0 ( 0.0) | 7 ( 5.1) |  |
| 1 | 6 ( 60.0) | 76 ( 55.5) |  |
| 2 | 4 ( 40.0) | 42 ( 30.7) |  |
| Miss | 0 ( 0.0) | 12 ( 8.8) |  |
| KNEE\_EXT\_SEL (%) |  |  | 0.371 |
| 0 | 0 ( 0.0) | 12 ( 8.8) |  |
| 1 | 3 ( 30.0) | 61 ( 44.5) |  |
| 2 | 5 ( 50.0) | 53 ( 38.7) |  |
| Miss | 2 ( 20.0) | 11 ( 8.0) |  |
| KNEE\_FLEX\_SEL (%) |  |  | 0.979 |
| 0 | 2 ( 20.0) | 21 ( 15.3) |  |
| 1 | 4 ( 40.0) | 59 ( 43.1) |  |
| 2 | 3 ( 30.0) | 45 ( 32.8) |  |
| Miss | 1 ( 10.0) | 12 ( 8.8) |  |
| PLANTFLEX\_SEL (%) |  |  | 0.442 |
| 0 | 3 ( 30.0) | 23 ( 16.8) |  |
| 1 | 5 ( 50.0) | 80 ( 58.4) |  |
| 2 | 0 ( 0.0) | 17 ( 12.4) |  |
| Miss | 2 ( 20.0) | 17 ( 12.4) |  |
| HIP\_ABD\_STR (%) |  |  | 0.768 |
| 2 | 1 ( 10.0) | 31 ( 22.6) |  |
| 3 | 5 ( 50.0) | 70 ( 51.1) |  |
| 4 | 3 ( 30.0) | 22 ( 16.1) |  |
| 5 | 0 ( 0.0) | 1 ( 0.7) |  |
| Miss | 1 ( 10.0) | 13 ( 9.5) |  |
| HIP\_EXT\_STR (%) |  |  | 0.792 |
| 1 | 0 ( 0.0) | 2 ( 1.5) |  |
| 2 | 2 ( 20.0) | 38 ( 27.7) |  |
| 3 | 4 ( 40.0) | 42 ( 30.7) |  |
| 4 | 2 ( 20.0) | 32 ( 23.4) |  |
| 5 | 0 ( 0.0) | 10 ( 7.3) |  |
| Miss | 2 ( 20.0) | 13 ( 9.5) |  |
| HIP\_FLEX\_STR (%) |  |  | 0.733 |
| 3 | 2 ( 20.0) | 25 ( 18.2) |  |
| 4 | 4 ( 40.0) | 60 ( 43.8) |  |
| 5 | 4 ( 40.0) | 40 ( 29.2) |  |
| Miss | 0 ( 0.0) | 12 ( 8.8) |  |
| KNEE\_EXT\_STR (%) |  |  | 0.652 |
| 2 | 0 ( 0.0) | 4 ( 2.9) |  |
| 3 | 2 ( 20.0) | 35 ( 25.5) |  |
| 4 | 1 ( 10.0) | 27 ( 19.7) |  |
| 5 | 5 ( 50.0) | 60 ( 43.8) |  |
| Miss | 2 ( 20.0) | 11 ( 8.0) |  |
| KNEE\_FLEX\_STR (%) |  |  | 0.904 |
| 2 | 1 ( 10.0) | 7 ( 5.1) |  |
| 3 | 3 ( 30.0) | 44 ( 32.1) |  |
| 4 | 3 ( 30.0) | 56 ( 40.9) |  |
| 5 | 2 ( 20.0) | 18 ( 13.1) |  |
| Miss | 1 ( 10.0) | 12 ( 8.8) |  |
| PLANTFLEX\_STR (%) |  |  | 0.846 |
| 0 | 0 ( 0.0) | 1 ( 0.7) |  |
| 1 | 1 ( 10.0) | 9 ( 6.6) |  |
| 2 | 6 ( 60.0) | 71 ( 51.8) |  |
| 3 | 1 ( 10.0) | 32 ( 23.4) |  |
| 4 | 0 ( 0.0) | 7 ( 5.1) |  |
| Miss | 2 ( 20.0) | 17 ( 12.4) |  |
| ADDUCTOR\_SPAS (%) |  |  | 0.329 |
| 1 | 7 ( 70.0) | 91 ( 66.4) |  |
| 2 | 2 ( 20.0) | 25 ( 18.2) |  |
| 3 | 0 ( 0.0) | 18 ( 13.1) |  |
| 4 | 1 ( 10.0) | 3 ( 2.2) |  |
| HAMSTRING\_SPAS (%) |  |  | 0.551 |
| 1 | 8 ( 80.0) | 96 ( 70.1) |  |
| 2 | 1 ( 10.0) | 33 ( 24.1) |  |
| 3 | 1 ( 10.0) | 8 ( 5.8) |  |
| HIP\_FLEX\_SPAS (%) |  |  | 0.794 |
| 1 | 8 ( 80.0) | 106 ( 77.4) |  |
| 2 | 2 ( 20.0) | 25 ( 18.2) |  |
| 3 | 0 ( 0.0) | 6 ( 4.4) |  |
| PLANTFLEX\_SPAS (%) |  |  | 0.835 |
| 1 | 5 ( 50.0) | 81 ( 59.1) |  |
| 2 | 3 ( 30.0) | 27 ( 19.7) |  |
| 3 | 2 ( 20.0) | 20 ( 14.6) |  |
| 4 | 0 ( 0.0) | 6 ( 4.4) |  |
| Miss | 0 ( 0.0) | 3 ( 2.2) |  |
| RECT\_FEM\_SPAS (%) |  |  | 0.922 |
| 1 | 5 ( 50.0) | 78 ( 56.9) |  |
| 2 | 3 ( 30.0) | 34 ( 24.8) |  |
| 3 | 1 ( 10.0) | 17 ( 12.4) |  |
| 4 | 1 ( 10.0) | 8 ( 5.8) |  |
| footsev (mean (SD)) | 1.32 (0.51) | 0.82 (0.39) | <0.001 |
| meanstaFooPrgdev (mean (SD)) | 23.13 (21.08) | 16.83 (13.86) | 0.184 |
| faqt (mean (SD)) | 54.19 (23.38) | 43.18 (23.26) | 0.151 |

#### Foot Physical Exam

|  | Control | Treated | p |
| --- | --- | --- | --- |
| n | 137 | 137 |  |
| NWB\_ARCH (%) |  |  | 0.966 |
| -1 | 87 (63.5) | 85 (62.0) |  |
| 0 | 35 (25.5) | 36 (26.3) |  |
| 1 | 15 (10.9) | 16 (11.7) |  |
| NWB\_FOREFT (%) |  |  | 0.002 |
| -2 | 82 (59.9) | 70 (51.1) |  |
| 0 | 41 (29.9) | 29 (21.2) |  |
| 2 | 14 (10.2) | 37 (27.0) |  |
| Miss | 0 ( 0.0) | 1 ( 0.7) |  |
| NWB\_FOREFT2 (%) |  |  | 0.990 |
| -1 | 2 ( 1.5) | 2 ( 1.5) |  |
| 0 | 104 (75.9) | 103 (75.2) |  |
| 1 | 31 (22.6) | 32 (23.4) |  |
| NWB\_FOREFT2\_SEVERITY (%) |  |  | 0.985 |
| 0 | 104 (75.9) | 103 (75.2) |  |
| 1 | 24 (17.5) | 26 (19.0) |  |
| 2 | 8 ( 5.8) | 7 ( 5.1) |  |
| 3 | 1 ( 0.7) | 1 ( 0.7) |  |
| NWB\_FOREFT\_SEVERITY (%) |  |  | 0.184 |
| 0 | 47 (34.3) | 48 (35.0) |  |
| 1 | 28 (20.4) | 36 (26.3) |  |
| 2 | 32 (23.4) | 36 (26.3) |  |
| 3 | 30 (21.9) | 17 (12.4) |  |
| NWB\_HINDFT (%) |  |  | 0.361 |
| -1 | 2 ( 1.5) | 3 ( 2.2) |  |
| 1 | 117 (85.4) | 123 (89.8) |  |
| 2 | 18 (13.1) | 11 ( 8.0) |  |
| NWB\_HINDFT\_SEVERITY (%) |  |  | 0.529 |
| 0 | 117 (85.4) | 123 (89.8) |  |
| 1 | 19 (13.9) | 13 ( 9.5) |  |
| 2 | 1 ( 0.7) | 1 ( 0.7) |  |
| NWB\_HIND\_EVER (%) |  |  | 0.930 |
| -1 | 57 (41.6) | 59 (43.1) |  |
| 0 | 69 (50.4) | 66 (48.2) |  |
| 1 | 11 ( 8.0) | 12 ( 8.8) |  |
| NWB\_HIND\_INVER (%) |  |  | 0.526 |
| -1 | 37 (27.0) | 29 (21.2) |  |
| 0 | 82 (59.9) | 89 (65.0) |  |
| 1 | 18 (13.1) | 19 (13.9) |  |
| NWB\_MID\_MOT (%) |  |  | 0.926 |
| -1 | 15 (10.9) | 17 (12.4) |  |
| 0 | 69 (50.4) | 67 (48.9) |  |
| 1 | 53 (38.7) | 53 (38.7) |  |
| NWB\_SUBTAL\_NEUT = 1 (%) | 129 (94.2) | 132 (96.4) | 0.570 |
| WB\_4FTPOS (%) |  |  | 0.461 |
| -2 | 51 (37.2) | 46 (33.6) |  |
| 0 | 75 (54.7) | 74 (54.0) |  |
| 2 | 11 ( 8.0) | 17 (12.4) |  |
| WB\_4FTPOS2 (%) |  |  | 0.966 |
| -1 | 56 (40.9) | 57 (41.6) |  |
| 0 | 44 (32.1) | 42 (30.7) |  |
| 1 | 37 (27.0) | 38 (27.7) |  |
| WB\_4FTPOS2\_SEVERITY (%) |  |  | 0.947 |
| 0 | 44 (32.1) | 42 (30.7) |  |
| 1 | 50 (36.5) | 48 (35.0) |  |
| 2 | 35 (25.5) | 37 (27.0) |  |
| 3 | 8 ( 5.8) | 10 ( 7.3) |  |
| WB\_4FTPOS\_SEVERITY (%) |  |  | 0.984 |
| 0 | 75 (54.7) | 74 (54.0) |  |
| 1 | 40 (29.2) | 39 (28.5) |  |
| 2 | 18 (13.1) | 19 (13.9) |  |
| 3 | 4 ( 2.9) | 5 ( 3.6) |  |
| WB\_FTPOS (%) |  |  | 0.205 |
| -4 | 8 ( 5.8) | 11 ( 8.0) |  |
| -3 | 89 (65.0) | 84 (61.3) |  |
| -2 | 13 ( 9.5) | 5 ( 3.6) |  |
| 0 | 13 ( 9.5) | 14 (10.2) |  |
| 2 | 5 ( 3.6) | 5 ( 3.6) |  |
| 3 | 9 ( 6.6) | 18 (13.1) |  |
| WB\_FTPOS\_SEVERITY (%) |  |  | 0.986 |
| 0 | 13 ( 9.5) | 14 (10.2) |  |
| 1 | 48 (35.0) | 49 (35.8) |  |
| 2 | 55 (40.1) | 55 (40.1) |  |
| 3 | 21 (15.3) | 19 (13.9) |  |
| WB\_MIDFT\_POS (%) |  |  | 0.327 |
| -1 | 10 ( 7.3) | 14 (10.2) |  |
| 0 | 22 (16.1) | 29 (21.2) |  |
| 1 | 105 (76.6) | 94 (68.6) |  |

### Kinematics Pre/Post x Treated/Control

### Effect size (Cohen’s D) for the chosen outcome variables

### Virtual Twins

- Build a BART model for the given outcome
- Create twin (synthetic counterfactual) for each observation
  - twin 1 has treatment of interest set to true for all observations
  - twin 0 has treatment of interest set to false for all observations
- Predict outcome for twin 1 and twin 0
- Compute difference in outcome: Effect = outcome(twin 1) - outcome(twin 0)

#### Note that we analyze two different samples using Virtual Twins

- Sample 1: **matched observations** from the direct matching analysis
- Sample 2: **all treated** limbs (effect of treatment on the treated)

### Model performance on outcome variables

### Plot the VT effect estimates

#### Matched subset

#### All treated limbs

### Bayesian causal forests (BCF)

The BCF method also uses a BART model, but in a different manner from the virtual twins approach. In BCF the outcome is explicitly split into a treatment effect (\(\tau\)) and a mean effect (\(\mu\)). The treatment effect only applies to treated limbs. Both treatment and mean effects are assumed to depend on the covariates, and, explicitly, on the propensity score. The latter corrects for regularization induced confounding

### Model performance

### Plot the BCF effect estimates

#### Matched subset

#### All treated limbs

### Compare Effect Size Estimates From All Models

Combine the estimated treatment effects from the three models (DM, VT, BCF) and two samples (matched, all-treated).
