## Supplementary material for "Estimating the Efficacy of Common Treatments in Children and Young Adults Diagnosed with Cerebral Palsy Using Three Machine Learning Algorithms": Detailed Results for Each of the 13 Treatments: Hams_Lengthening.html

### Results

This document contains details related to the estimation of treatment effects at four levels for three different models:

- Direct Matching (DM)
- Virtual Twins (VT)
- Bayesian Causal Forests (BCF)

##### Treatment of interest

Hams Lengthening

##### Outcomes

- **Anatomy & Physiology**: POP\_ANG\_UNI
- **Gait Parameter**: minswiKneFlx
- **Global Gait**: GDI
- **Function**: faqt

#### Direct Matching

We use the *designmatch* package to do direct matching. Each treatment has custom matching parameters:

- Distance (pairwise distance between matches)
  - this also includes the propensity score as a caliper
  - weighting on the caliper adjusts how close propensity matches
- Near Fine
  - mostly useful for matching interval surgery
  - occasionally used to match distributions of factors (e.g. strength)
- Moment
  - ensures mean distributions match between groups

##### Variables for distance matching

GDI, HIP\_ABD\_0, POP\_ANG\_UNI, foHipFlx, foKneFlx, icHipFlx, icKneFlx, maxHipFlx, maxKneFlx, maxstaHipFlx, maxstaKneFlx, maxswiHipFlx, maxswiKneFlx, meanHipFlx, meanKneFlx, meanstaHipFlx, meanstaKneFlx, meanswiHipFlx, meanswiKneFlx, minHipFlx, minKneFlx, minstaHipFlx, minstaKneFlx, minswiHipFlx, minswiKneFlx, romHipFlx, romKneFlx, romstaHipFlx, romstaKneFlx, romswiHipFlx, romswiKneFlx, velKneFlx, faqt, AGE, NDspeed, NDsteplen

##### Variables for moment matching

HIP\_ABD\_0, AGE, NDspeed, NDsteplen, POP\_ANG\_UNI, minswiKneFlx, GDI, faqt

##### Variables for near-fine matching

Neural Rhizotomy, Neurotoxin Injection, Adductor Release, Gastroc Soleus Lengthening, Psoas Release, Patellar Advance, Femoral Derotation Osteotomy, Tibial Derotation Osteotomy, DFEO, gmfcs

Building the matching problem… GLPK optimizer is open… Finding the optimal matches… Optimal matches found

##### Matched subset of treated and control limbs

Number of Matching Pairs = 76 out of 86

#### Covariate balance

#### Propensity score balance

#### Prior and interval treatment balance

#### Covariate distributions

This is the same data in the balance plots, but **complete** and in explicit tabular form

For foot surgeries, an additional table showing foot physical exam data is included

##### Matched pairs

|  | Control | Treated | p |
| --- | --- | --- | --- |
| n | 76 | 76 |  |
| AGE (mean (SD)) | 11.62 (2.53) | 11.28 (2.92) | 0.449 |
| GDI (mean (SD)) | 71.01 (9.71) | 71.40 (11.20) | 0.820 |
| NDspeed (mean (SD)) | 0.31 (0.09) | 0.31 (0.08) | 0.555 |
| NDsteplen (mean (SD)) | 0.60 (0.13) | 0.59 (0.12) | 0.627 |
| footoff (mean (SD)) | 0.63 (0.06) | 0.62 (0.04) | 0.385 |
| oppfootoff (mean (SD)) | 0.13 (0.06) | 0.13 (0.04) | 0.780 |
| oppfootcontact (mean (SD)) | 0.49 (0.04) | 0.49 (0.03) | 0.644 |
| ANTEVERSION (mean (SD)) | 39.08 (16.50) | 33.59 (19.85) | 0.066 |
| BIMAL (mean (SD)) | 17.00 (13.42) | 18.83 (12.71) | 0.390 |
| HIP\_INT\_ROT (mean (SD)) | 62.25 (12.45) | 57.14 (16.93) | 0.036 |
| HIP\_EXT\_ROT (mean (SD)) | 31.30 (14.48) | 34.42 (14.60) | 0.188 |
| HIP\_ABD\_0 (mean (SD)) | 28.46 (9.29) | 29.30 (10.01) | 0.592 |
| HIP\_ABD\_90 (mean (SD)) | 58.26 (11.85) | 61.20 (14.87) | 0.180 |
| HIP\_FLEX (mean (SD)) | 127.88 (11.82) | 131.07 (9.28) | 0.067 |
| HIP\_EXT (mean (SD)) | 4.58 (10.88) | 1.74 (10.52) | 0.104 |
| KNEE\_EXT (mean (SD)) | 3.49 (8.48) | 2.82 (6.90) | 0.593 |
| KNEE\_FLEX (mean (SD)) | 134.28 (9.32) | 134.38 (8.51) | 0.942 |
| POP\_ANG\_UNI (mean (SD)) | 63.45 (11.97) | 65.07 (12.03) | 0.407 |
| ANK\_DORS\_0 (mean (SD)) | -2.32 (11.09) | -1.96 (9.09) | 0.829 |
| ANK\_DORS\_90 (mean (SD)) | 10.90 (11.42) | 10.24 (10.02) | 0.706 |
| foAnkDor (mean (SD)) | -3.41 (11.83) | -4.62 (14.32) | 0.571 |
| SEX (mean (SD)) | 0.66 (0.48) | 0.64 (0.48) | 0.866 |
| dx = Cerebral palsy (%) | 76 (100.0) | 76 (100.0) | NA |
| dxmod (%) |  |  | 0.686 |
| Diplegia | 38 ( 50.0) | 42 ( 55.3) |  |
| Hemiplegia | 8 ( 10.5) | 7 ( 9.2) |  |
| Hemiplegia type II | 0 ( 0.0) | 2 ( 2.6) |  |
| Hemiplegia type III | 1 ( 1.3) | 0 ( 0.0) |  |
| Hemiplegia type IV | 1 ( 1.3) | 1 ( 1.3) |  |
| Quadriplegia | 14 ( 18.4) | 14 ( 18.4) |  |
| Triplegia | 14 ( 18.4) | 10 ( 13.2) |  |
| dxside (%) |  |  | 0.164 |
| Asymmetrical L | 13 ( 17.1) | 15 ( 19.7) |  |
| Asymmetrical R | 14 ( 18.4) | 12 ( 15.8) |  |
| Bilateral | 26 ( 34.2) | 29 ( 38.2) |  |
| Left | 3 ( 3.9) | 10 ( 13.2) |  |
| Right | 14 ( 18.4) | 8 ( 10.5) |  |
| Unknown | 6 ( 7.9) | 2 ( 2.6) |  |
| affected (mean (SD)) | 0.87 (0.34) | 0.87 (0.34) | 1.000 |
| gmfcs (%) |  |  | 0.648 |
| 1 | 18 ( 23.7) | 17 ( 22.4) |  |
| 2 | 21 ( 27.6) | 24 ( 31.6) |  |
| 3 | 13 ( 17.1) | 11 ( 14.5) |  |
| 4 | 2 ( 2.6) | 0 ( 0.0) |  |
| Miss | 22 ( 28.9) | 24 ( 31.6) |  |
| HIP\_ABD\_SEL (%) |  |  | 0.617 |
| 0 | 11 ( 14.5) | 7 ( 9.2) |  |
| 1 | 36 ( 47.4) | 33 ( 43.4) |  |
| 2 | 23 ( 30.3) | 29 ( 38.2) |  |
| Miss | 6 ( 7.9) | 7 ( 9.2) |  |
| HIP\_EXT\_SEL (%) |  |  | 0.243 |
| 0 | 9 ( 11.8) | 7 ( 9.2) |  |
| 1 | 37 ( 48.7) | 27 ( 35.5) |  |
| 2 | 24 ( 31.6) | 31 ( 40.8) |  |
| Miss | 6 ( 7.9) | 11 ( 14.5) |  |
| HIP\_FLEX\_SEL (%) |  |  | 0.918 |
| 0 | 1 ( 1.3) | 2 ( 2.6) |  |
| 1 | 44 ( 57.9) | 44 ( 57.9) |  |
| 2 | 26 ( 34.2) | 24 ( 31.6) |  |
| Miss | 5 ( 6.6) | 6 ( 7.9) |  |
| KNEE\_EXT\_SEL (%) |  |  | 0.172 |
| 0 | 6 ( 7.9) | 10 ( 13.2) |  |
| 1 | 40 ( 52.6) | 28 ( 36.8) |  |
| 2 | 27 ( 35.5) | 31 ( 40.8) |  |
| Miss | 3 ( 3.9) | 7 ( 9.2) |  |
| KNEE\_FLEX\_SEL (%) |  |  | 0.235 |
| 0 | 9 ( 11.8) | 9 ( 11.8) |  |
| 1 | 49 ( 64.5) | 38 ( 50.0) |  |
| 2 | 13 ( 17.1) | 23 ( 30.3) |  |
| Miss | 5 ( 6.6) | 6 ( 7.9) |  |
| PLANTFLEX\_SEL (%) |  |  | 0.227 |
| 0 | 21 ( 27.6) | 25 ( 32.9) |  |
| 1 | 38 ( 50.0) | 27 ( 35.5) |  |
| 2 | 9 ( 11.8) | 9 ( 11.8) |  |
| Miss | 8 ( 10.5) | 15 ( 19.7) |  |
| HIP\_ABD\_STR (%) |  |  | 0.605 |
| 0 | 1 ( 1.3) | 0 ( 0.0) |  |
| 1 | 0 ( 0.0) | 1 ( 1.3) |  |
| 2 | 23 ( 30.3) | 17 ( 22.4) |  |
| 3 | 32 ( 42.1) | 38 ( 50.0) |  |
| 4 | 11 ( 14.5) | 12 ( 15.8) |  |
| 5 | 3 ( 3.9) | 1 ( 1.3) |  |
| Miss | 6 ( 7.9) | 7 ( 9.2) |  |
| HIP\_EXT\_STR (%) |  |  | 0.466 |
| 0 | 1 ( 1.3) | 0 ( 0.0) |  |
| 2 | 24 ( 31.6) | 17 ( 22.4) |  |
| 3 | 22 ( 28.9) | 19 ( 25.0) |  |
| 4 | 17 ( 22.4) | 22 ( 28.9) |  |
| 5 | 6 ( 7.9) | 7 ( 9.2) |  |
| Miss | 6 ( 7.9) | 11 ( 14.5) |  |
| HIP\_FLEX\_STR (%) |  |  | 0.175 |
| 3 | 16 ( 21.1) | 16 ( 21.1) |  |
| 4 | 33 ( 43.4) | 21 ( 27.6) |  |
| 5 | 22 ( 28.9) | 33 ( 43.4) |  |
| Miss | 5 ( 6.6) | 6 ( 7.9) |  |
| KNEE\_EXT\_STR (%) |  |  | 0.249 |
| 2 | 3 ( 3.9) | 0 ( 0.0) |  |
| 3 | 35 ( 46.1) | 32 ( 42.1) |  |
| 4 | 12 ( 15.8) | 16 ( 21.1) |  |
| 5 | 23 ( 30.3) | 21 ( 27.6) |  |
| Miss | 3 ( 3.9) | 7 ( 9.2) |  |
| KNEE\_FLEX\_STR (%) |  |  | 0.079 |
| 2 | 6 ( 7.9) | 0 ( 0.0) |  |
| 3 | 29 ( 38.2) | 24 ( 31.6) |  |
| 4 | 30 ( 39.5) | 41 ( 53.9) |  |
| 5 | 6 ( 7.9) | 5 ( 6.6) |  |
| Miss | 5 ( 6.6) | 6 ( 7.9) |  |
| PLANTFLEX\_STR (%) |  |  | 0.430 |
| 0 | 1 ( 1.3) | 2 ( 2.6) |  |
| 1 | 7 ( 9.2) | 5 ( 6.6) |  |
| 2 | 41 ( 53.9) | 35 ( 46.1) |  |
| 3 | 15 ( 19.7) | 15 ( 19.7) |  |
| 4 | 4 ( 5.3) | 2 ( 2.6) |  |
| 5 | 0 ( 0.0) | 2 ( 2.6) |  |
| Miss | 8 ( 10.5) | 15 ( 19.7) |  |
| ADDUCTOR\_SPAS (%) |  |  | 0.462 |
| 1 | 44 ( 57.9) | 52 ( 68.4) |  |
| 2 | 11 ( 14.5) | 11 ( 14.5) |  |
| 3 | 14 ( 18.4) | 9 ( 11.8) |  |
| 4 | 7 ( 9.2) | 4 ( 5.3) |  |
| HAMSTRING\_SPAS (%) |  |  | 0.069 |
| 1 | 36 ( 47.4) | 48 ( 63.2) |  |
| 2 | 32 ( 42.1) | 19 ( 25.0) |  |
| 3 | 8 ( 10.5) | 7 ( 9.2) |  |
| 4 | 0 ( 0.0) | 2 ( 2.6) |  |
| HIP\_FLEX\_SPAS (%) |  |  | 0.037 |
| 1 | 49 ( 64.5) | 55 ( 72.4) |  |
| 2 | 27 ( 35.5) | 17 ( 22.4) |  |
| 3 | 0 ( 0.0) | 4 ( 5.3) |  |
| PLANTFLEX\_SPAS (%) |  |  | 0.433 |
| 1 | 32 ( 42.1) | 30 ( 39.5) |  |
| 2 | 21 ( 27.6) | 24 ( 31.6) |  |
| 3 | 16 ( 21.1) | 16 ( 21.1) |  |
| 4 | 7 ( 9.2) | 3 ( 3.9) |  |
| 5 | 0 ( 0.0) | 1 ( 1.3) |  |
| Miss | 0 ( 0.0) | 2 ( 2.6) |  |
| RECT\_FEM\_SPAS (%) |  |  | 0.096 |
| 1 | 39 ( 51.3) | 48 ( 63.2) |  |
| 2 | 19 ( 25.0) | 18 ( 23.7) |  |
| 3 | 13 ( 17.1) | 10 ( 13.2) |  |
| 4 | 5 ( 6.6) | 0 ( 0.0) |  |
| minswiKneFlx (mean (SD)) | 31.02 (11.67) | 32.39 (10.44) | 0.449 |
| faqt (mean (SD)) | 42.92 (23.80) | 44.68 (27.36) | 0.672 |

##### Unmatched treated vs. Matched treated

This comparison is useful for understanding possible bias from failing to match some treated observations

|  | Unmatched | Matched | p |
| --- | --- | --- | --- |
| n | 10 | 76 |  |
| AGE (mean (SD)) | 9.51 (2.73) | 11.28 (2.92) | 0.073 |
| GDI (mean (SD)) | 68.71 (11.96) | 71.40 (11.20) | 0.481 |
| NDspeed (mean (SD)) | 0.25 (0.09) | 0.31 (0.08) | 0.028 |
| NDsteplen (mean (SD)) | 0.54 (0.10) | 0.59 (0.12) | 0.203 |
| footoff (mean (SD)) | 0.64 (0.06) | 0.62 (0.04) | 0.252 |
| oppfootoff (mean (SD)) | 0.15 (0.06) | 0.13 (0.04) | 0.189 |
| oppfootcontact (mean (SD)) | 0.48 (0.05) | 0.49 (0.03) | 0.687 |
| ANTEVERSION (mean (SD)) | 56.60 (15.69) | 33.59 (19.85) | 0.001 |
| BIMAL (mean (SD)) | 15.30 (16.96) | 18.83 (12.71) | 0.430 |
| HIP\_INT\_ROT (mean (SD)) | 75.10 (10.70) | 57.14 (16.93) | 0.002 |
| HIP\_EXT\_ROT (mean (SD)) | 25.30 (12.19) | 34.42 (14.60) | 0.062 |
| HIP\_ABD\_0 (mean (SD)) | 28.70 (10.86) | 29.30 (10.01) | 0.860 |
| HIP\_ABD\_90 (mean (SD)) | 56.50 (14.35) | 61.20 (14.87) | 0.348 |
| HIP\_FLEX (mean (SD)) | 131.00 (8.43) | 131.07 (9.28) | 0.983 |
| HIP\_EXT (mean (SD)) | -1.90 (13.35) | 1.74 (10.52) | 0.322 |
| KNEE\_EXT (mean (SD)) | 0.25 (5.46) | 2.82 (6.90) | 0.261 |
| KNEE\_FLEX (mean (SD)) | 133.40 (3.66) | 134.38 (8.51) | 0.721 |
| POP\_ANG\_UNI (mean (SD)) | 70.20 (12.84) | 65.07 (12.03) | 0.212 |
| ANK\_DORS\_0 (mean (SD)) | 3.20 (12.58) | -1.96 (9.09) | 0.111 |
| ANK\_DORS\_90 (mean (SD)) | 18.25 (11.79) | 10.24 (10.02) | 0.022 |
| foAnkDor (mean (SD)) | -10.93 (20.74) | -4.62 (14.32) | 0.219 |
| SEX (mean (SD)) | 0.80 (0.42) | 0.64 (0.48) | 0.335 |
| dx = Cerebral palsy (%) | 10 (100.0) | 76 (100.0) | NA |
| dxmod (%) |  |  | 0.953 |
| Diplegia | 5 ( 50.0) | 42 ( 55.3) |  |
| Hemiplegia | 1 ( 10.0) | 7 ( 9.2) |  |
| Hemiplegia type II | 0 ( 0.0) | 2 ( 2.6) |  |
| Hemiplegia type IV | 0 ( 0.0) | 1 ( 1.3) |  |
| Quadriplegia | 3 ( 30.0) | 14 ( 18.4) |  |
| Triplegia | 1 ( 10.0) | 10 ( 13.2) |  |
| dxside (%) |  |  | 0.267 |
| Asymmetrical L | 1 ( 10.0) | 15 ( 19.7) |  |
| Asymmetrical R | 0 ( 0.0) | 12 ( 15.8) |  |
| Bilateral | 7 ( 70.0) | 29 ( 38.2) |  |
| Left | 0 ( 0.0) | 10 ( 13.2) |  |
| Right | 2 ( 20.0) | 8 ( 10.5) |  |
| Unknown | 0 ( 0.0) | 2 ( 2.6) |  |
| affected (mean (SD)) | 0.90 (0.32) | 0.87 (0.34) | 0.782 |
| gmfcs (%) |  |  | 0.106 |
| 1 | 0 ( 0.0) | 17 ( 22.4) |  |
| 2 | 4 ( 40.0) | 24 ( 31.6) |  |
| 3 | 4 ( 40.0) | 11 ( 14.5) |  |
| Miss | 2 ( 20.0) | 24 ( 31.6) |  |
| HIP\_ABD\_SEL (%) |  |  | 0.080 |
| 0 | 3 ( 30.0) | 7 ( 9.2) |  |
| 1 | 1 ( 10.0) | 33 ( 43.4) |  |
| 2 | 4 ( 40.0) | 29 ( 38.2) |  |
| Miss | 2 ( 20.0) | 7 ( 9.2) |  |
| HIP\_EXT\_SEL (%) |  |  | 0.920 |
| 0 | 1 ( 10.0) | 7 ( 9.2) |  |
| 1 | 4 ( 40.0) | 27 ( 35.5) |  |
| 2 | 3 ( 30.0) | 31 ( 40.8) |  |
| Miss | 2 ( 20.0) | 11 ( 14.5) |  |
| HIP\_FLEX\_SEL (%) |  |  | 0.349 |
| 0 | 0 ( 0.0) | 2 ( 2.6) |  |
| 1 | 7 ( 70.0) | 44 ( 57.9) |  |
| 2 | 1 ( 10.0) | 24 ( 31.6) |  |
| Miss | 2 ( 20.0) | 6 ( 7.9) |  |
| KNEE\_EXT\_SEL (%) |  |  | 0.064 |
| 0 | 4 ( 40.0) | 10 ( 13.2) |  |
| 1 | 3 ( 30.0) | 28 ( 36.8) |  |
| 2 | 1 ( 10.0) | 31 ( 40.8) |  |
| Miss | 2 ( 20.0) | 7 ( 9.2) |  |
| KNEE\_FLEX\_SEL (%) |  |  | 0.168 |
| 0 | 2 ( 20.0) | 9 ( 11.8) |  |
| 1 | 6 ( 60.0) | 38 ( 50.0) |  |
| 2 | 0 ( 0.0) | 23 ( 30.3) |  |
| Miss | 2 ( 20.0) | 6 ( 7.9) |  |
| PLANTFLEX\_SEL (%) |  |  | 0.572 |
| 0 | 5 ( 50.0) | 25 ( 32.9) |  |
| 1 | 3 ( 30.0) | 27 ( 35.5) |  |
| 2 | 0 ( 0.0) | 9 ( 11.8) |  |
| Miss | 2 ( 20.0) | 15 ( 19.7) |  |
| HIP\_ABD\_STR (%) |  |  | 0.438 |
| 1 | 1 ( 10.0) | 1 ( 1.3) |  |
| 2 | 1 ( 10.0) | 17 ( 22.4) |  |
| 3 | 5 ( 50.0) | 38 ( 50.0) |  |
| 4 | 1 ( 10.0) | 12 ( 15.8) |  |
| 5 | 0 ( 0.0) | 1 ( 1.3) |  |
| Miss | 2 ( 20.0) | 7 ( 9.2) |  |
| HIP\_EXT\_STR (%) |  |  | 0.798 |
| 2 | 3 ( 30.0) | 17 ( 22.4) |  |
| 3 | 3 ( 30.0) | 19 ( 25.0) |  |
| 4 | 2 ( 20.0) | 22 ( 28.9) |  |
| 5 | 0 ( 0.0) | 7 ( 9.2) |  |
| Miss | 2 ( 20.0) | 11 ( 14.5) |  |
| HIP\_FLEX\_STR (%) |  |  | 0.204 |
| 3 | 1 ( 10.0) | 16 ( 21.1) |  |
| 4 | 5 ( 50.0) | 21 ( 27.6) |  |
| 5 | 2 ( 20.0) | 33 ( 43.4) |  |
| Miss | 2 ( 20.0) | 6 ( 7.9) |  |
| KNEE\_EXT\_STR (%) |  |  | 0.612 |
| 3 | 3 ( 30.0) | 32 ( 42.1) |  |
| 4 | 3 ( 30.0) | 16 ( 21.1) |  |
| 5 | 2 ( 20.0) | 21 ( 27.6) |  |
| Miss | 2 ( 20.0) | 7 ( 9.2) |  |
| KNEE\_FLEX\_STR (%) |  |  | 0.105 |
| 3 | 6 ( 60.0) | 24 ( 31.6) |  |
| 4 | 2 ( 20.0) | 41 ( 53.9) |  |
| 5 | 0 ( 0.0) | 5 ( 6.6) |  |
| Miss | 2 ( 20.0) | 6 ( 7.9) |  |
| PLANTFLEX\_STR (%) |  |  | 0.368 |
| 0 | 0 ( 0.0) | 2 ( 2.6) |  |
| 1 | 3 ( 30.0) | 5 ( 6.6) |  |
| 2 | 3 ( 30.0) | 35 ( 46.1) |  |
| 3 | 2 ( 20.0) | 15 ( 19.7) |  |
| 4 | 0 ( 0.0) | 2 ( 2.6) |  |
| 5 | 0 ( 0.0) | 2 ( 2.6) |  |
| Miss | 2 ( 20.0) | 15 ( 19.7) |  |
| ADDUCTOR\_SPAS (%) |  |  | 0.190 |
| 1 | 4 ( 40.0) | 52 ( 68.4) |  |
| 2 | 4 ( 40.0) | 11 ( 14.5) |  |
| 3 | 1 ( 10.0) | 9 ( 11.8) |  |
| 4 | 1 ( 10.0) | 4 ( 5.3) |  |
| HAMSTRING\_SPAS (%) |  |  | 0.946 |
| 1 | 6 ( 60.0) | 48 ( 63.2) |  |
| 2 | 3 ( 30.0) | 19 ( 25.0) |  |
| 3 | 1 ( 10.0) | 7 ( 9.2) |  |
| 4 | 0 ( 0.0) | 2 ( 2.6) |  |
| HIP\_FLEX\_SPAS (%) |  |  | 0.182 |
| 1 | 7 ( 70.0) | 55 ( 72.4) |  |
| 2 | 1 ( 10.0) | 17 ( 22.4) |  |
| 3 | 2 ( 20.0) | 4 ( 5.3) |  |
| PLANTFLEX\_SPAS (%) |  |  | 0.898 |
| 1 | 3 ( 30.0) | 30 ( 39.5) |  |
| 2 | 3 ( 30.0) | 24 ( 31.6) |  |
| 3 | 3 ( 30.0) | 16 ( 21.1) |  |
| 4 | 1 ( 10.0) | 3 ( 3.9) |  |
| 5 | 0 ( 0.0) | 1 ( 1.3) |  |
| Miss | 0 ( 0.0) | 2 ( 2.6) |  |
| RECT\_FEM\_SPAS (%) |  |  | 0.893 |
| 1 | 6 ( 60.0) | 48 ( 63.2) |  |
| 2 | 3 ( 30.0) | 18 ( 23.7) |  |
| 3 | 1 ( 10.0) | 10 ( 13.2) |  |
| minswiKneFlx (mean (SD)) | 34.62 (9.27) | 32.39 (10.44) | 0.522 |
| faqt (mean (SD)) | 27.90 (13.78) | 44.68 (27.36) | 0.061 |

#### Kinematics Pre/Post x Treated/Control

#### Effect size (Cohen’s D) for the chosen outcome variables

#### Virtual Twins

- Build a BART model for the given outcome
- Create twin (synthetic counterfactual) for each observation
  - twin 1 has treatment of interest set to true for all observations
  - twin 0 has treatment of interest set to false for all observations
- Predict outcome for twin 1 and twin 0
- Compute difference in outcome: Effect = outcome(twin 1) - outcome(twin 0)

##### Note that we analyze two different samples using Virtual Twins

- Sample 1: **matched observations** from the direct matching analysis
- Sample 2: **all treated** limbs (effect of treatment on the treated)

#### Model performance on outcome variables

#### Plot the VT effect estimates

##### Matched subset

##### All treated limbs

#### Bayesian causal forests (BCF)

The BCF method also uses a BART model, but in a different manner from the virtual twins approach. In BCF the outcome is explicitly split into a treatment effect (\(\tau\)) and a mean effect (\(\mu\)). The treatment effect only applies to treated limbs. Both treatment and mean effects are assumed to depend on the covariates, and, explicitly, on the propensity score. The latter corrects for regularization induced confounding

#### Model performance

#### Plot the BCF effect estimates

##### Matched subset

##### All treated limbs

#### Compare Effect Size Estimates From All Models

Combine the estimated treatment effects from the three models (DM, VT, BCF) and two samples (matched, all-treated).
